# WHITE-Net: White matter HyperIntensities Tissue Extraction using deep learning Network

**DOI:** 10.1101/2025.01.09.25320242

**Authors:** Camille Cathala, Vincent Roduit, Fábio Carneiro, Manon Heffernan, Quentin Vanderbecq, Olivier Colliot, Ferath Kherif, Jean-Philippe Thiran, Aurélie Bussy, Bogdan Draganski

**Author notes:** Equal contribution (Aurélie Bussy, Bogdan Draganski).

## Abstract

White matter hyperintensities (WMH) are a hallmark of cerebral small vessel disease, a highly prevalent condition in aging, making their accurate detection in large-scale magnetic resonance imaging (MRI) studies critically important. Currently available semi-automated and automated segmentation methods are frequently limited by sensitivity to variations in imaging conditions and computational constraints, restricting their applicability across diverse acquisition settings. We present WHITE-Net, a deep learning-based framework for automated WMH segmentation built on a 3D ResUNet architecture and trained on multisite MRI data encompassing substantial variability in scanner vendors, acquisition protocols, spatial resolution, and image characteristics. Evaluated across three independent datasets - BrainLaus, ADNI, and WMH Challenge, WHITE-Net achieves high segmentation accuracy and ranks among the top-performing methods across datasets, enabling a comprehensive assessment of generalizability in varied imaging environments. WHITE-Net maintains stable performance in a wide spectrum of lesion loads and demonstrates a favorable balance between precision and recall, effectively limiting false positive detections - a common limitation of existing approaches. Beyond accuracy, WHITE-Net requires no parameter tuning and offers fast inference times, making it well-suited for deployment in large-scale neuroimaging studies. The obtained results highlight the value of training on diverse multisite data for improving generalizability and position WHITE-Net as a reliable, scalable tool for automated WMH segmentation in computational anatomy research.

## 1 Introduction

White Matter Hyperintensities (WMH) are areas of abnormal signal intensity on diagnostic magnetic resonance imaging (MRI) protocols sensitive to tissue water content, such as fluid attenuated inversion recovery (FLAIR). Their etiology varies depending on the clinical context: WMHs of presumed vascular origin are strongly associated with cardiovascular risk factors and cerebral amyloid angiopathy (Aljondi et al., 2018; de Bresser et al., 2018; Fuhrmann et al., 2019; Ghaznawi et al., 2018; Kuller et al., 2004; Trofimova et al., 2023), while in migraine (Kruit et al., 2009; Seneviratne et al., 2013), and multiple sclerosis (Fazekas et al., 1999; Filippi et al., 2019; Rovira & León, 2008) the underlying processes are inflammatory or partially unknown; and in leukodystrophies they reflect hereditary or acquired disorders of myelin maintenance. WMHs of vascular origin are strongly associated with aging and represent a common incidental finding in the general population, with prevalence rising beyond the age of 60 years. They are a hallmark feature of cerebral small vessel disease (cSVD), which is a major contributor to vascular cognitive impairment (Coenen et al., 2022; Jiménez-Balado et al., 2022), and dementia (Lee et al., 2016; Mortamais et al., 2013).

The importance of automated, accurate detection of WMHs complementary to the neuroradiological diagnostic expertise, is well recognized in both clinical and research context. The sheer volume of data acquired in large-scale MRI studies precludes manual lesion delineation, making automated WMH segmentation indispensable. Despite its well-known limitations in terms of time investment and inter-rater variability (Commowick et al., 2018; Grimaud et al., 1996), manual lesion annotation remains the gold standard in neuroimaging. Currently available automated WMH segmentation algorithms span a broad spectrum from intensity-based approaches (Gaser et al., 2022; Schmidt et al., 2012), to machine learning (Griffanti et al., 2016) and deep learning methods (Li et al., 2018b; Park et al., 2021).

Intensity-based methods rely on predefined or automatically determined thresholds to identify WMH based on voxel intensity and morphology. While straightforward, these approaches are often limited by their sensitivity to noise and imaging protocols variability. Machine learning approaches overcome this by training classifiers on intensity and spatial features, better capturing lesion variability, although generalizability across heterogenous datasets remains a challenge. Deep learning models, in particular convolutional neural networks (CNNs), automatically extract complex features from imaging data, improving WMH segmentation performance, but require large training datasets and considerable computational resources (Rachmadi et al., 2018). Despite their strong performance, CNN-based methods are frequently affected by variability in scanner hardware, image acquisition protocols, and image quality, which can limit their robustness and generalizability in multisite settings (Heinen et al., 2019).

Existing WMH segmentation methods offer clear advantages in accuracy and time efficiency relative to manual annotation. However, most algorithms are trained on data comprising well-defined conditions in clinical populations, such as multiple sclerosis (Schmidt, 2017; Schmidt et al., 2012), stroke or neurodegeneration (Griffanti et al., 2016; Li et al., 2018b; Park et al., 2021), which reduces their generalizability to epidemiological MRI datasets. The emergence of large-scale studies in the community-dwelling population incorporating brain MRI, such as the UK Biobank (Sudlow et al., 2015) targeting 100 ‘000 acquisitions, highlights the pressing need for accurate and automated WMH segmentation tools. Robust and reliable segmentation across cohorts characterized by a wide age range, image quality variability, and a spectrum of aging-associated pathologies is essential to ensure the validity of computational anatomy studies at scale.

Current evaluations of WMH segmentation methods predominantly rely on global overlap metrics such as the Dice coefficient, which may obscure important differences in segmentation behavior (Maier-Hein et al., 2018). In particular, methods achieving similar Dice scores can exhibit markedly different error profiles with respect to false positive detections and missed lesions (Seghier, 2024). A more comprehensive evaluation framework encompassing lesion-level metrics and analysis across varying lesion loads and imaging conditions, is therefore necessary to more fully characterize method performance.

Against this background, we present WHITE-Net (White matter HyperIntensities Tissue Extraction Network), a fast, user-friendly, and generalizable deep learning algorithm for WMH segmentation based on a 3D ResUNet architecture. Unlike many existing approaches, WHITE-Net is trained on multisite MRI datasets encompassing substantial variability in scanner vendors, acquisition protocols, spatial resolution, and image characteristics, with the aim of improving robustness to domain shifts and enhancing generalizability across heterogeneous datasets. Performance is evaluated across multiple independent datasets and benchmarked against established methods using both global and lesion-level metrics. Beyond overall accuracy, we systematically assess robustness across distribution shifts, performance stratified by lesion load, lesion size, and scanner characteristics; and differences in segmentation error profiles across methods.

Taken together, the results demonstrate that WHITE-Net achieves accurate and robust WMH segmentation across heterogeneous imaging datasets, with strong generalizability across varying lesion loads, scanner types, and acquisition protocols. It maintains a favorable balance between precision and recall, effectively limiting false positive detections. These characteristics makes WHITE-Net particularly well-suited for large-scale epidemiological neuroimaging studies, where robustness to imaging variability and reliable lesion detection are essential.

## 2 Methods

### 2.1 Datasets

We used three MRI datasets with distinct demographic characteristics, imaging protocols, and clinico-epidemiological profiles: BrainLaus, the WMH Segmentation Challenge, and the Alzheimer’s Disease Neuroimaging Initiative (ADNI). The BrainLaus cohort consists of community-dwelling individuals, whereas ADNI and the WMH Challenge datasets include older participants with neurodegenerative and cerebrovascular pathologies. Together, these datasets provide substantial variability in scanner hardware and acquisition protocols, enabling evaluation of model robustness and generalization.

Throughout the manuscript, unless explicitly stated otherwise, the terms in-distribution (ID) and out-of-distribution (OOD) refer specifically to the relationship between the evaluation datasets and the training distribution of WHITE-Net. Thus, BrainLaus and the WMH Challenge training distribution are considered ID for WHITE-Net, whereas the WMH Challenge test data acquired with scanners not represented during WHITE-Net training and the ADNI test data are considered OOD. These labels should not be interpreted as universal properties of the datasets or as describing the training distribution of the other comparison methods, whose models were trained on different datasets or, in the case of methods such as BIANCA, trained specifically in the present study. For pretrained comparison methods, their own original training datasets and corresponding distributional coverage are described in Section 2.10.

#### 2.1.1 BrainLaus dataset

The BrainLaus dataset originates from the CoLaus*|*PsycoLaus cohort, a population-based study investigating associations between cardiovascular risk factors and mental health (Firmann et al., 2008; Preisig et al., 2009). A subset of 163 participants was selected based on image quality and Fazekas score distribution (mean age: 59.65 years; range: 19.78–92.83; 56.6% female). The dataset was split into training (n=81) and testing (n=82) sets.

#### 2.1.2 WMH challenge dataset

The WMH Segmentation Challenge dataset (H. Kuijf et al., 2022) includes individuals with neurodegenerative and cerebrovascular conditions acquired under heterogeneous imaging protocols. The training set comprises 60 scans acquired on three MRI scanners (3T Philips Achieva, 3T Siemens TrioTim, and 3T GE Signa HDx), while the test set includes 110 individuals acquired on five scanners, including 3T Philips Achieva, 3T Siemens TrioTim, 3T GE Signa HDx, 3T Philips Ingenuity, and 1.5T GE Signa HDxt.

#### 2.1.3 ADNI dataset

The Alzheimer’s Disease Neuroimaging Initiative (ADNI) is a multicenter longitudinal study aimed at developing imaging biomarkers (Jack et al., 2008; Mueller et al., 2005). It includes cognitively normal individuals (NC), patients with mild cognitive impairment (MCI), and Alzheimer’s disease, providing a broad spectrum of neurodegenerative phenotypes. Here, a subset of 115 scans with available FLAIR MRI was selected (Vanderbecq et al., 2020) for the training set including heterogeneous acquisitions from multiple 3T scanners (Siemens, GE, and Philips), while the test set contains 32 scans acquired on previously unseen scanners, allowing evaluation of cross-scanner generalization.

### 2.2 MRI acquisition

#### 2.2.1 BrainLaus dataset

MRI data were acquired on a 3T whole-body system (Magnetom Prisma - Siemens, Erlangen Germany), using a 64-channel radiofrequency receive head coil and a body coil for transmission. FLAIR acquisition parameters are provided in Table 1.

**Table 1:**
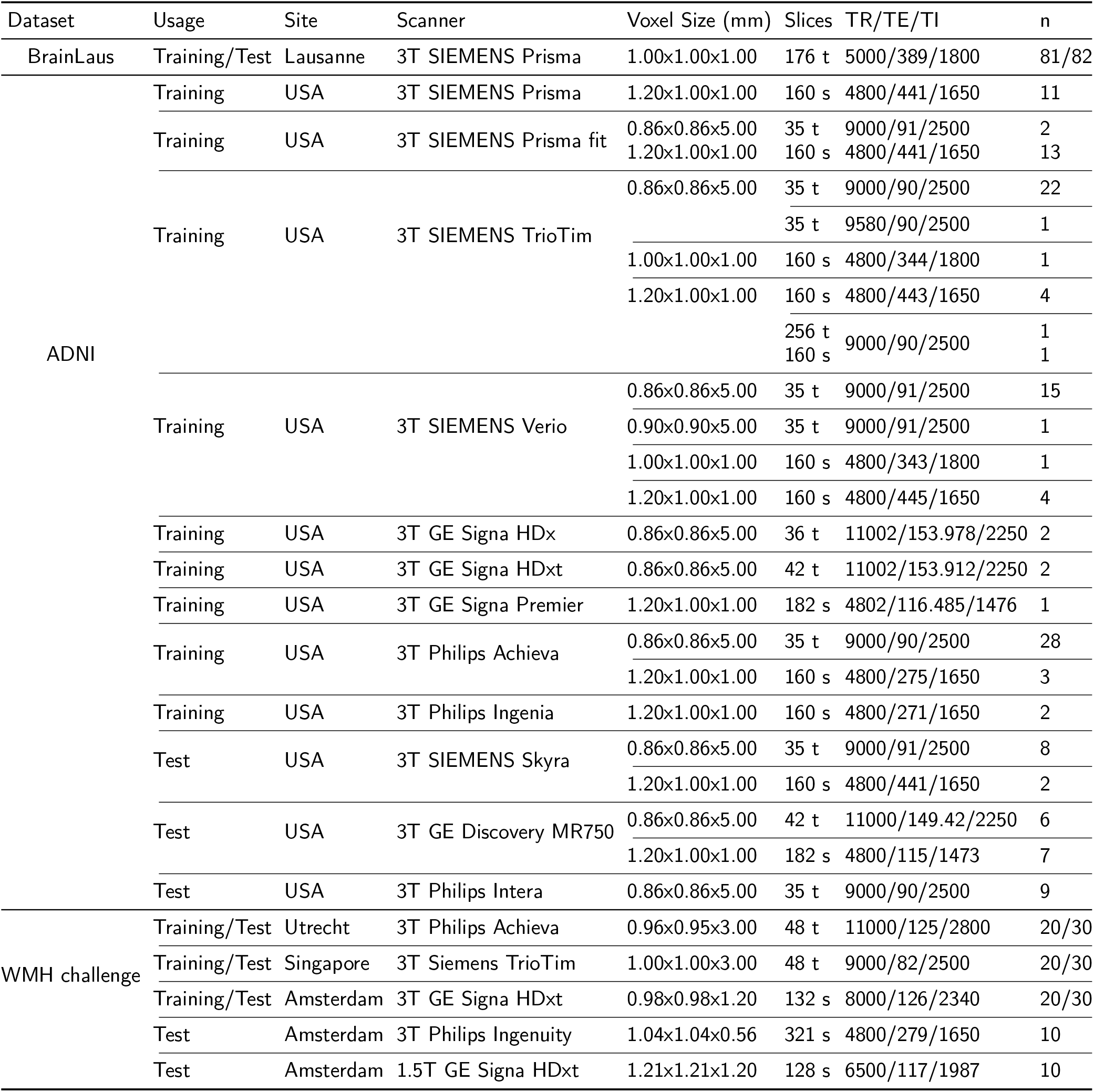
Table summarizing MRI sequence parameters across all included datasets (BrainLaus, ADNI, WMH Challenge), detailing scanner- and site-specific acquisition settings, along with the distribution of MRIs used for training and testing, including sample sizes (n) per group. Slices are reported with orientation: t = axial (transversal), s = sagittal.

#### 2.2.2 WMH challenge dataset

MRI data were acquired across five scanners from different vendors and field strengths. Acquisition parameters and scanner details are summarized in Table 1 (Boomsma et al., 2017; van Veluw et al., 2015).

#### 2.2.3 ADNI dataset

ADNI MRI data were acquired across multiple sites using harmonized protocols on 3T scanners from Siemens, GE, and Philips (Jack et al., 2008). Detailed acquisition parameters are provided in Table 1.

### 2.3 MRI preprocessing

All MRI data were preprocessed using SPM12-based (www.fil.ion.ucl.ac.uk/spm) CAT12 toolbox (Gaser et al., 2022) in MATLAB R2021 (MathWorks, Sherborn, MA, USA). Images were denoised using a spatial adaptive non-local means filter (Manjón et al., 2009) and corrected for intensity non-uniformity using bias field correction.

In BrainLaus, quality control was performed based on visual inspection for motion artifacts (Wood & Henkelman, 1985). A motion score ranging from 1 (minimal motion) to 4 (severe motion) with 0.5 increments was assigned, and only scans with a score below 2.5 were retained.

In BrainLaus, WMH severity was additionally assessed using the Fazekas scale (Fazekas et al., 1987), evaluating periventricular and deep white matter lesions (0-3 each), resulting in a combined score ranging from 0 to 6. Participants were selected to ensure balanced representation across Fazekas scores.

### 2.4 Image standardization

All images underwent skull-stripping using SPM12s unified segmentation, followed by min-max intensity normalization. Data were resampled and zero-padded to a fixed size of 192 *×* 176 *×* 160 voxels. If resampling was required, corresponding WM and WMH masks were transformed back to original space for analysis.

### 2.5 Manual annotation

#### 2.5.1 BrainLaus dataset

Manual WMH annotation was performed on FLAIR images using the MINC-tool Display software. Lesions were manually delineated slice-by-slice by trained raters (CC and AB), with simultaneous visualization in all three orthogonal planes to ensure spatial consistency. Prior to independent annotation, both raters underwent a training on approximately 20 data sets to harmonize lesion definition criteria. Raters were blinded to Fazekas scores during segmentation to avoid bias. Participants were selected based on image quality to span the full spectrum of WMH burden (Fazekas 0–6), ensuring representation of both small and large lesions and approximately balanced sampling across Fazekas levels. Selection was not based on age or sex. Demographic characteristics across Fazekas levels are reported in Supplementary Table 1. Segmentation followed standard criteria for WMH identification on FLAIR images, comprising hyperintense regions within the white matter while excluding non-pathological signal changes and anatomically ambiguous regions. Specifically, lesions in subcortical grey matter and the brainstem were not annotated, and thin periventricular hyperintense rims (one voxel in thickness) were excluded to avoid lesion overestimation. All segmentations were subsequently reviewed and validated by an experienced neurologist (FC), ensuring anatomical accuracy and consistency across participants.

#### 2.5.2 WMH challenge dataset

WMH annotations were generated by four expert raters per scan (H. J. Kuijf et al., 2019). Final binary masks were obtained by thresholding the consensus annotations, with voxels reaching at least 50% rater agreement labeled as WMH.

#### 2.5.3 ADNI dataset

WMH annotation was carried out by a trained radiology resident using ITK-SNAP (Yushkevich et al., 2006) and following standardized guidelines, excluding non-pathological signal changes and anatomically ambiguous regions to ensure consistency across subjects (Vanderbecq et al., 2020).

### 2.6 Data Augmentation

To improve model robustness and generalization across heterogeneous MRI acquisitions, we applied during training on-the-fly data augmentation using the TorchIO framework (Pérez-García et al., 2021). The augmentation combined spatial transformations and intensity-based perturbations to simulate realistic variability of MRI data (Supplementary Figure 2).

Spatial augmentations included random right-left flipping, affine transformations, and elastic deformations, enabling the model to learn invariance to anatomical orientation, scaling, and non-linear deformations. Acquisition-related variability was simulated through random anisotropy to mimic differences in voxel spatial resolution, and through motion, ghosting, and spike artifacts that reproduce common MRI acquisition imperfections.

Intensity-based augmentations included bias field simulation to account for coil inhomogeneity and gamma transformations that model contrast variability. Rician noise was additionally introduced to realistically emulate MRI magnitude noise, implemented by adding Gaussian noise to both real and imaginary signal components of the signal and calculating the resulting magnitude data.

To further increase data diversity, each training sample is replicated four times, generating one original and three augmented versions per subject. Augmentations were applied probabilistically, ensuring a wide range of transformations while preserving anatomical plausibility and were disabled during validation.

### 2.7 Network architecture

The proposed architecture is a ResUNet (Ronneberger et al., 2015), variant of the standard ResUNet (Diakogiannis et al., 2020) that integrates residual learning to enhance image segmentation performance. The model takes FLAIR images as inputs and is trained to produce binary WMH masks.

The architecture comprises an encoder and a decoder path (Supplementary Figure 3). The encoder progressively reduces spatial information while increasing feature dimensionality through a series of convolutions layers, with Rectified Linear Unit (ReLU) activations and max-pooling. Within each block, a residual skip connection adds the block input directly to its output, mitigating the “vanishing gradient” problem and improving the network’s ability to learn complex features representations.

The decoder reconstructs feature and spatial resolution information through deconvolutions and concatenation with high-resolution features from the corresponding encoder levels via skip connections. Dropout layers are incorporated to prevent overfitting. The encoder begins with 16 kernels, doubling at each downsampling step. The decoder mirrors this pattern in reverse, reducing the kernel count as spatial resolution is restored.

### 2.8 Implementation information

The network is implemented in Python3 using the PyTorch framework (Paszke et al., 2019). The model is trained on one NVIDIA RTX A4000 GPU with 16GB RAM. The optimization process is performed with an Adam optimizer, batch size = 2, learning rate = 1*e^−^*^4^ for 300 epochs. 20 % of the training set, randomly selected, is used as the validation set. The model weights are saved each time the validation metric increases. Early stopping is used if the validation loss did not decrease for 20 epochs. The model is optimized using a variant of the Dice loss (Milletari et al., 2016) as follows:

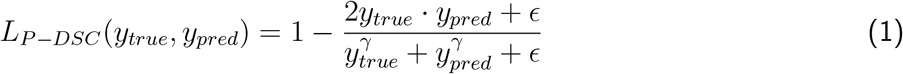

with *γ* = 2.The power *γ* increases the sensitivity to difference in prediction. It amplifies the differences, therefore the loss largely penalizes misclassification.

During inference, an adaptive thresholding strategy is applied to refine the final lesion segmentation from the predicted probability map. First, an initial binary mask is obtained using a fixed threshold (0.5) to estimate lesion load, defined as the number of voxels above this threshold. Cases are categorized based on lesion load as low (<300 voxels), medium (300–5000 voxels), or high (>5000 voxels), which determines the adaptive decision threshold (0.8, 0.7, and 0.5, respectively) to account for variability in disease burden. Connected component analysis is then performed, and only components with at least 5 voxels whose mean probability exceeds the adaptive threshold are retained in the final segmentation mask.

### 2.9 Performance evaluation

#### 2.9.1 Voxel-level metrics

- The Dice Similarity coefficient (DSC) is used to evaluate the spatial overlap between the manually annotated WMHs and the generated WMH mask using the following equation. 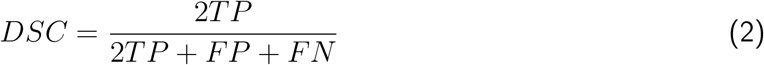 where TP, FP, and FN denote the number of true positive, false positive, and false negative voxels, respectively. If both the prediction and the ground truth are empty, DSC is set to 1.
- The absolute volume difference (AVD) between manual WMH segmentation and algorithm output is defined as 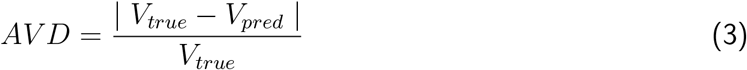 If the manual segmentation contains no lesions, the AVD is set to 0 if the predicted mask is empty and 1 if the reverse is true.
- The 95th percentile Hausdorff distance (HD95) measures the boundary discrepancy between the manual and automated segmentations. 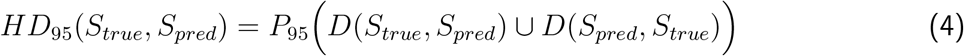 where *D*(*S_true_, S_pred_*) denotes the set of shortest Euclidean distances from each boundary voxel in *S_true_* to the closest voxel in *S_pred_*, and vice versa. Hausdorff distance is defined only if the manual segmentation and the predicted mask contain lesions.

#### 2.9.2 Lesion matching procedure

For each subject, ground-truth and predicted WMH masks were decomposed into individual lesion clusters using connected-component analysis (26-connectivity). Candidate correspondences between ground-truth and predicted lesions were quantified using the intersection-over-union (IoU), and an optimal one-to-one assignment between ground-truth and predicted lesions was obtained using the Hungarian algorithm (linear assignment problem), maximizing the total IoU across matched pairs. Assigned pairs with IoU *≥* 0.1 were classified as true positives (TP). Assigned pairs below this threshold, along with any unmatched ground-truth and predicted lesions, were classified as false negatives (FN) and false positives (FP), respectively. Note that because matching is enforced as one-to-one, a predicted lesion spatially overlapping multiple ground-truth lesions (or vice versa) is matched to only one counterpart, with the remaining lesion(s) counted as FN or FP.

#### 2.9.3 Lesion-level metrics

- WMH volume was calculated by multiplying the number of voxels in the manually segmented WMH mask by the voxel volume derived from the NIfTI image header, yielding total WMH volume in mm^3^. For analyses involving total WMH volume, values were log_10_-transformed as log_10_(WMH volume+1) to reduce skewness and accommodate scans with zero WMH volume.
- Lesion-wise recall evaluates the proportion of ground-truth lesions correctly detected, i.e., matched to a predicted lesion according to the procedure described in Section 2.9.2. 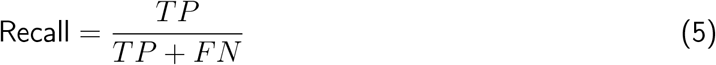
- Lesion-wise precision evaluates the proportion of predicted lesions that correspond to a true lesion. 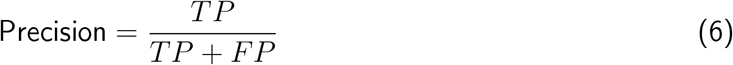
- The lesion-wise F1-score is defined as the harmonic mean of lesion-wise recall and precision. 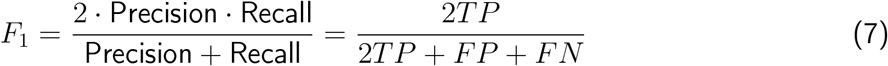 Note that this expression is mathematically identical to the Dice coefficient (Eq. 1): the two metrics differ only in the level at which TP, FP, and FN are defined: voxel-wise for DSC, lesion-wise (Section 2.9.2) for *F*_1_.

### 2.10 Performance comparison with other WMH segmentation tools

WHITE-Net was compared with a range of publicly available WMH segmentation tools and frameworks, encompassing three methodological categories: (i) classical or probabilistic approaches that do not require supervised training on the target dataset, (ii) generic machine-learning frameworks that require training on the target dataset, and (iii) pretrained WMH-specific models developed and trained on independent datasets. In this third category, performance reflects both the underlying segmentation architecture and the characteristics of the data and training procedure used by the original developers.

#### Classical and probabilistic methods

The following methods were applied using their publicly available implementations and recommended settings, without retraining on the present datasets:

- **LPA**: Lesion Prediction Algorithm (Schmidt, 2017) from the lesion segmentation toolbox (LST) for SPM12. LPA is a supervised machine-learning approach based on logistic regression and was originally developed for the detection of white matter lesions in FLAIR images.
- **LGA**: Lesion Growth Algorithm (Schmidt et al., 2012) from the lesion segmentation toolbox (LST) for SPM12. LGA uses a probabilistic model and region-growing procedure to identify WMH based on FLAIR and T1-weighted images.
- **CAT12** (Gaser et al., 2022): The Computational Anatomy Toolbox based for SPM12. Its WMH detection procedure uses tissue probability maps and image intensity information to identify voxels and clusters compatible with WMH.
- **SamSeg** (Cerri et al., 2021; Puonti et al., 2016): A parametric Bayesian whole-brain and lesion segmentation method, implemented within the FreeSurfer framework. SamSeg estimates anatomical structures and lesion classes using probabilistic models and does not require training on the target dataset.

#### Generic machine-learning framework

- **BIANCA** (Griffanti et al., 2016): The Brain Intensity AbNormality Classification Algorithm, implemented in FSL. BIANCA uses a k-nearest-neighbour classifier and therefore requires training examples from the target dataset. In the present study, BIANCA was trained using the same training set as WHITE-Net and was evaluated using two input configurations: FLAIR only (BIANCA_1_) and FLAIR together with T1-weighted images (BIANCA_2_). Following systematic evaluation, a probability threshold of 0.1 was selected for BIANCA.

#### Pretrained WMH-specific deep-learning models

The following methods were evaluated using pretrained models released by their respective authors. None of these models was retrained or fine-tuned on the BrainLaus, ADNI, or WMH Challenge datasets used in the present study.

- **PGS** (Park et al., 2021): The PGS model, originally developed for the 2017 MICCAI WMH Segmentation Challenge (H. J. Kuijf et al., 2019). PGS is an ensemble of U-Net models with multi-scale highlighting of foreground regions. The model was trained on the 60-subject training set of the WMH Segmentation Challenge (presented in Section 2.1), which comprised paired T1-weighted and FLAIR MRI acquired at three institutions using five scanners. The remaining 110 subjects formed the challenge test set and were not available for model training (H. J. Kuijf et al., 2019). The publicly released pretrained model was used directly in the present study.
- **MARS-WMH**_1_ **(MD-GRU)** (Andermatt et al., 2018; Gesierich et al., 2025): This model is one of two pretrained deep-learning pipelines released as part of MARS-WMH. The model was trained by the MARS-WMH authors on a heterogeneous dataset comprising 304 subjects from three cohorts: ADNI (*n* = 244), RUNDMC (*n* = 38), and VASCAMY (*n* = 22). The model was trained using multimodal MRI data including FLAIR and T1-weighted images. The training data were selected to cover a broad range of WMH volumes and MRI hardware, including different scanner models and acquisition protocols. WMH reference masks were generated semi-automatically and subsequently manually corrected by an expert rater according to STRIVE criteria; lacunes were excluded. This pretrained MARS-WMH model is based on a multi-dimensional gated recurrent unit (MD-GRU) architecture. The pretrained model was applied without retraining or fine-tuning.
- **MARS-WMH**_2_ **(nnU-Net)** (Gesierich et al., 2025; Isensee et al., 2020): The nnU-Net-based model from MARS-WMH. Importantly, this model was trained within the MARS-WMH framework using the same heterogeneous training dataset and reference annotations as the MARS-WMH_1_ model. The publicly released MARS-WMH_2_ model was applied without retraining or fine-tuning.
- **segcsvd** (Gibson et al., 2024): A convolutional neural network-based WMH segmentation method developed for heterogeneous clinical cohorts. The model was developed using 947 3T FLAIR scans from seven studies, encompassing heterogeneous patient populations, imaging parameters, and WMH burden. The training data included cohorts with Alzheimer’s disease, vascular cognitive impairment, aphasia, sleep apnea, carotid stenosis, and healthy controls. The method uses FLAIR together with anatomical information derived from SynthSeg in a two-stage segmentation pipeline. The publicly released model was applied without retraining or fine-tuning on the present datasets.
- **SYSU-MEDIA2** (Li et al., 2018a): A U-Net-like fully convolutional neural network using an ensemble of three networks trained with different initializations and data augmentation. The publicly available implementation was successfully executed on our data but failed to produce meaningful WMH segmentation outputs on BrainLaus and was therefore excluded from the quantitative comparisons.

All algorithms were applied to the test sets of the BrainLaus, ADNI, and WMH Challenge datasets. LPA, LGA, CAT12, and SamSeg were applied using their publicly available implementations and recommended settings. For PGS, MARS-WMH_1_, MARS-WMH_2_, and segcsvd, we used the pretrained models provided by the respective authors, without retraining or fine-tuning on our datasets. Therefore, the pretrained deep-learning comparisons assess the performance of the publicly available models as complete WMH segmentation tools, including their original training data, preprocessing, model architecture, and training procedures. They do not constitute controlled comparisons of the underlying neural-network architectures, since the models were trained on different datasets and with different procedures.

Pairwise statistical comparisons between segmentation methods were based on subject-level Dice scores. For each dataset, paired permutation tests were conducted across all method pairs by computing the mean difference in Dice scores and assessing significance using 10,000 random sign permutations. P-values were adjusted for multiple comparisons using the Benjamini-Hochberg false discovery rate correction (*α* = 0.05). Results are summarized as adjusted p-value matrices in Supplementary Figure 4.

## 3 Results

### 3.1 Dataset composition and imaging characteristics

Dataset composition and input MRI characteristics for BrainLaus, ADNI, and WMH Challenge cohorts are illustrated in (Figure 1). Sample sizes are comparable across datasets (BrainLaus: n=163; ADNI n= 147; WMH Challenge n= 170). Age distributions differ markedly: ADNI and WMH Challenge are confined to older participants with a relatively narrow age range, whereas BrainLaus encompasses a wider distribution including younger individuals. In BrainLaus, WMH burden increased monotonically with Fazekas score, confirming the expected alignment between visual rating and lesion volume.

**Figure 1:**
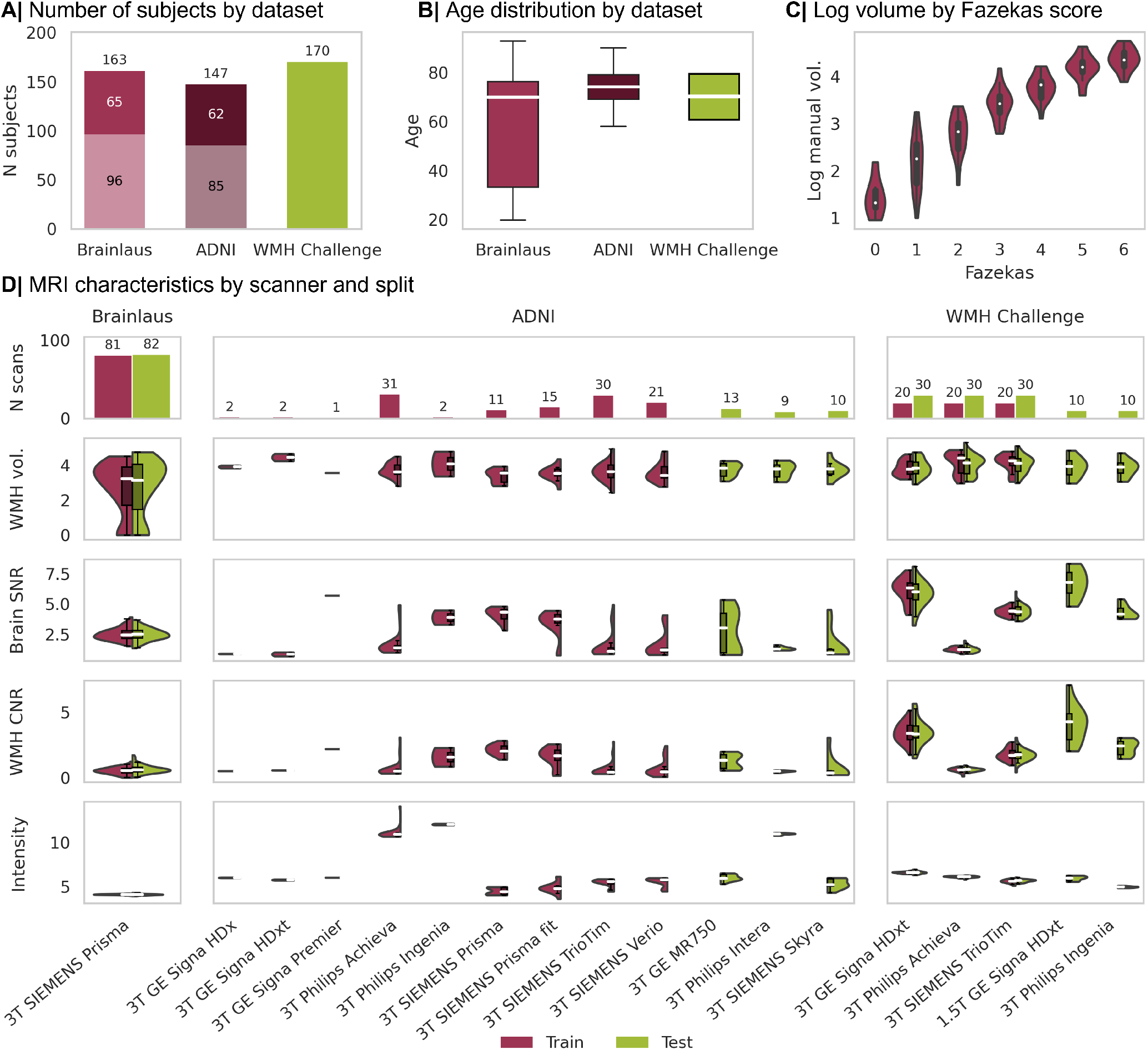
Dataset composition and input image characteristics across cohorts. (A) Number of subjects in each dataset, with dark and light shades representing males and females, respectively, when sex information was available. (B) Age distribution for BrainLaus, ADNI and WMH Challenge, the reported mean and standard deviation are shown. (C) Distribution of log_10_(manual WMH volume + 1) across Fazekas scores in BrainLaus. (D) Scanner-specific characteristics of the input data for each dataset, shown separately for training and test sets. The top row shows the number of scans per scanner, followed by the distribution of WMH volume, brain signal-to-noise ratio (SNR), WMH-to-NAWM contrast-to-noise ratio (CNR), and median brain intensity.

There was substantial heterogeneity in MRI acquisition across datasets and scanners (Table 1). BrainLaus data were acquired on a single scanner with balanced train/test splits, while ADNI and WMH Challenge data acquisition was distributed across multiple sites with variable representation across splits. Differences in WMH volume distributions and image quality metrics, including brain SNR, WMH-to-NAWM CNR, and voxel intensity distributions were the parameters reflecting the diversity of imaging conditions arising from differences in scanners and acquisition protocols.

Fifty-two MRIs from five out-of-distribution sites are included in the evaluation - two from the WMH Challenge and three from ADNI, representing scanner vendors and magnetic field strengths not encountered during training. These sites also show variability in key image characteristics, including brain SNR, WMH-to-NAWM CNR, and the voxel intensity range.

### 3.2 Overall performance of segmentation methods

To provide an overall assessment of the methods across datasets, we used the Dice coefficient as a global metric, as it captures both false positives and false negatives. Figure 2A shows the ranking of methods based on their mean Dice score per dataset. Detailed mean Dice scores and standard deviations for each method are reported in Table 2. For BrainLaus, WHITE-Net ranked first, followed by MARS-WMH_2_, SamSeg, and MARS-WMH_1_, all achieving Dice scores above 0.5. Conversely, segcsvd, LGA, and PGS showed moderate performance (0.35-0.5), while LPA, BIANCA, and CAT12 performed poorly (Dice < 0.3). For ADNI, PGS ranked first, followed by WHITE-Net, segcsvd, and MARS-WMH_1_, with intermediate performance from BIANCA, SamSeg, LPA, and MARS-WMH_2_. LGA and CAT12 showed the lowest performance. For the WMH Challenge dataset, PGS ranked first, followed by WHITE-Net. The overall ranking is similar to ADNI, although some differences are observed, with MARS-WMH_2_ performing better and SamSeg worse. Other metrics such as lesion recall, precision and F1-score, absolute volume difference (AVD) and Hausdorff distance, as well as execution time are presented in Table 2.

**Figure 2:**
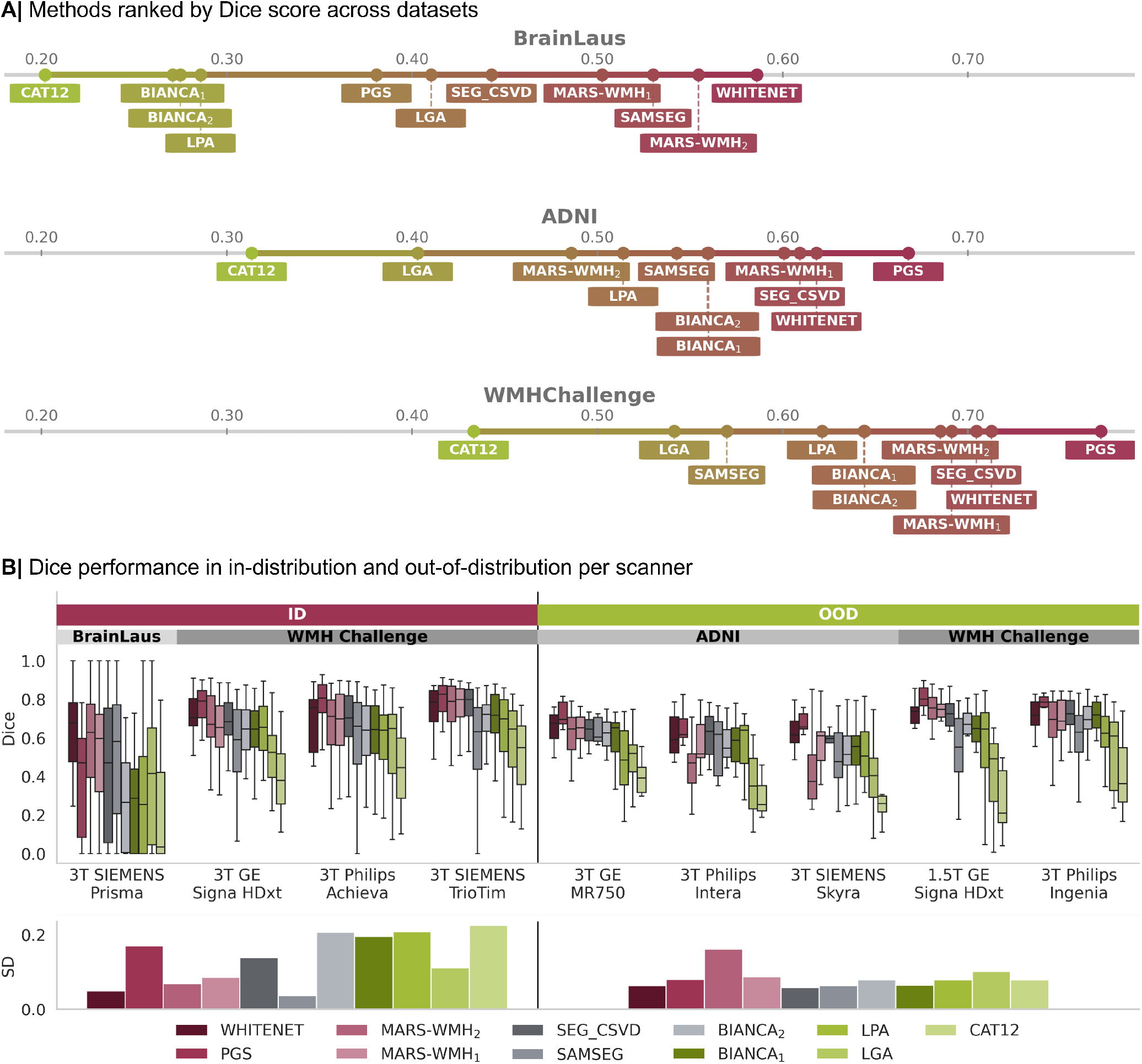
(A) Methods ranked by mean Dice score within each dataset. (B) Dice performance stratified by method and scanner, distinguishing in-distribution (ID) and out-of-distribution (OOD) settings with respect to the WHITE-Net training distribution. Standard deviation of Dice scores across sites for ID and OOD conditions, reflecting robustness to scanner variability.

**Table 2:** Performance comparison across test set of the 3 datasets. Mean *±* SD of the metrics are reported. Bold results outline the best performing algorithms. Higher is better for Dice, Recall, Precision, and F1. Lower is better for HD95 and AVD.

| Algorithm | Dataset | Dice | Lesion Recall | Lesion Precision | Lesion F1 | HD95 | AVD | Execution time |
| --- | --- | --- | --- | --- | --- | --- | --- | --- |
| LPA | BrainLaus | 0.29±0.26 | 0.17±0.21 | 0.42±0.35 | 0.20±0.18 | 28.00±22.85 | 0.65±1.21 | 38s |
| | ADNI | 0.52±0.16 | 0.47±0.26 | 0.64±0.21 | 0.46±0.19 | 16.69±11.93 | 1.07±1.80 | |
| | WMHChallenge | 0.62±0.19 | 0.37±0.19 | 0.68±0.21 | 0.42±0.13 | 15.00±9.82 | 0.67±1.17 | |
| LGA | BrainLaus | 0.41±0.34 | 0.22±0.31 | 0.53±0.37 | 0.26±0.30 | 22.21±18.23 | 0.75±2.89 | 128s |
| | ADNI | 0.41±0.18 | 0.19±0.10 | 0.74±0.21 | 0.27±0.09 | 25.75±14.31 | 0.62±0.35 | |
| | WMHChallenge | 0.54±0.21 | 0.19±0.11 | 0.74±0.17 | 0.29±0.12 | 20.64±14.69 | 2.17±18.66 | |
| CAT12 | BrainLaus | 0.20±0.26 | 0.39±0.27 | 0.27±0.29 | 0.25±0.22 | 34.63±30.47 | 310±1064 | 976s |
| | ADNI | 0.32±0.14 | 0.35±0.10 | 0.48±0.18 | 0.38±0.10 | 14.25±7.57 | 0.45±0.36 | |
| | WMHChallenge | 0.43±0.19 | 0.37±0.13 | 0.58±0.19 | 0.42±0.11 | 13.96±9.09 | 0.38±0.23 | |
| SamSeg | BrainLaus | 0.53±0.35 | 0.33±0.37 | 0.74±0.34 | 0.39±0.35 | 17.96±17.89 | 0.49±0.63 | 626s |
| | ADNI | 0.54±0.17 | 0.25±0.14 | 0.80±0.18 | 0.36±0.15 | 21.72±18.08 | 0.41±0.45 | |
| | WMHChallenge | 0.57±0.23 | 0.20±0.14 | 0.77±0.22 | 0.29±0.16 | 21.36±17.02 | 0.40±0.32 | |
| BIANCA<sub>1</sub> | BrainLaus | 0.27±0.23 | 0.22±0.22 | 0.39±0.35 | 0.20±0.15 | 28.75±27.09 | 1.02±1.94 | 603s |
| | ADNI | 0.56±0.14 | 0.42±0.19 | 0.70±0.20 | 0.47±0.15 | 14.01±9.69 | 0.36±0.30 | |
| | WMHChallenge | 0.64±0.16 | 0.39±0.17 | 0.70±0.23 | 0.45±0.15 | 12.50±9.28 | 0.37±0.55 | |
| BIANCA<sub>2</sub> | BrainLaus | 0.27±0.23 | 0.22±0.22 | 0.42±0.37 | 0.22±0.17 | 26.98±24.96 | 0.77±1.30 | 687s |
| | ADNI | 0.56±0.15 | 0.46±0.19 | 0.67±0.20 | 0.49±0.14 | 12.46±8.97 | 0.37±0.39 | |
| | WMHChallenge | 0.64±0.17 | 0.46±0.18 | 0.63±0.24 | 0.47±0.14 | 12.21±9.71 | 0.44±0.77 | |
| MARS-WMH<sub>1</sub> | BrainLaus | 0.50±0.30 | 0.59±0.32 | 0.62±0.37 | 0.58±0.32 | 18.46±27.72 | 0.45±1.20 | 169s |
| | ADNI | 0.60±0.12 | 0.61±0.18 | 0.75±0.16 | 0.64±0.11 | 14.00±12.14 | 0.33±0.28 | |
| | WMHChallenge | 0.69±0.16 | 0.75±0.14 | 0.64±0.17 | 0.67±0.13 | 11.77±11.63 | 0.40±0.48 | |
| MARS-WMH<sub>2</sub> | BrainLaus | 0.55±0.30 | **0.62±0.30** | 0.70±0.36 | 0.64±0.31 | **14.70±22.71** | 0.36±0.69 | 111s |
| | ADNI | 0.50±0.19 | 0.41±0.16 | **0.92±0.11** | 0.54±0.14 | 21.19±15.81 | 0.49±0.28 | |
| | WMHChallenge | 0.68±0.19 | 0.63±0.18 | 0.83±0.12 | 0.69±0.13 | 12.34±16.64 | 0.27±0.23 | |
| PGS | BrainLaus | 0.38±0.27 | 0.62±0.34 | 0.46±0.33 | 0.49±0.32 | 24.24±30.65 | 1.22±3.17 | 52s |
| | ADNI | **0.67±0.12** | **0.70±0.14** | 0.71±0.14 | **0.70±0.13** | 10.06±9.97 | **0.22±0.25** | |
| | WMHChallenge | **0.77±0.13** | **0.78±0.15** | 0.78±0.12 | **0.77±0.11** | **7.04±9.73** | **0.21±0.21** | |
| segcsvd | BrainLaus | 0.44±0.33 | 0.45±0.29 | 0.52±0.37 | 0.45±0.29 | 21.65±28.64 | 1.12±2.61 | 367s |
| | ADNI | 0.61±0.10 | 0.53±0.18 | 0.83±0.13 | 0.62±0.13 | **9.11±7.56** | 0.47±0.42 | |
| | WMHChallenge | 0.70±0.14 | 0.47±0.15 | 0.88±0.17 | 0.59±0.14 | 8.35±8.99 | 0.33±0.47 | |
| WHITE-Net | BrainLaus | **0.59±0.30** | 0.53±0.30 | **0.79±0.34** | **0.61±0.30** | 15.10±22.12 | **0.31±0.43** | **28s** |
| | ADNI | 0.62±0.12 | 0.43±0.14 | 0.89±0.13 | 0.56±0.12 | 11.84±9.15 | 0.24±0.25 | |
| | WMHChallenge | 0.71±0.13 | 0.46±0.15 | **0.90±0.11** | 0.59±0.13 | 9.43±9.09 | 0.22±0.20 | |

To assess whether differences in Dice scores between methods were statistically significant, we performed pairwise comparisons for each dataset (Supplementary figure 4). For BrainLaus, WHITE-Net, MARS-WMH_2_, and SamSeg showed similar performance and were superior to the other methods. For ADNI and the WMH Challenge datasets, PGS achieved significantly higher Dice scores than all other methods. In ADNI, WHITE-Net, segcsvd, and MARS-WMH_1_ performed similarly, whereas in the WMH Challenge dataset, WHITE-Net and segcsvd showed comparable performance and outperformed MARS-WMH_1_. Full pairwise comparison results are provided in the matrices for completeness.

Performance also varied across scanners (Figure 2B), with substantial differences in Dice scores depending on the acquisition site. WHITE-Net and SamSeg showed relatively consistent performance across scanners, although SamSeg achieved lower Dice scores overall. In contrast, MARS-WMH_2_, PGS, BIANCA, CAT12, and LPA exhibited greater variability, indicating reduced robustness to changes in imaging conditions. This was supported by the standard deviation of Dice scores across sites in both ID and OOD settings, where WHITE-Net demonstrated consistently low variability, while several other methods showed larger performance fluctuations across sites.

Performance was additionally evaluated under artificial transformations applied to the test set. Certain image artifacts had a stronger negative impact on Dice scores, particularly motion and anisotropy, whereas others had only a limited effect (e.g., gamma and spike; Supplementary Figure 7). Isotropic scans were less sensitive to artifact-induced degradation, while highly anisotropic images showed greater sensitivity, which increased with artifact severity.

### 3.3 Visual comparison of segmentation methods

Figure 3 shows qualitative WMH segmentation results for one representative subject from each dataset. The manual reference is displayed in pink, while the predictions of each method are shown in green.

**Figure 3:**
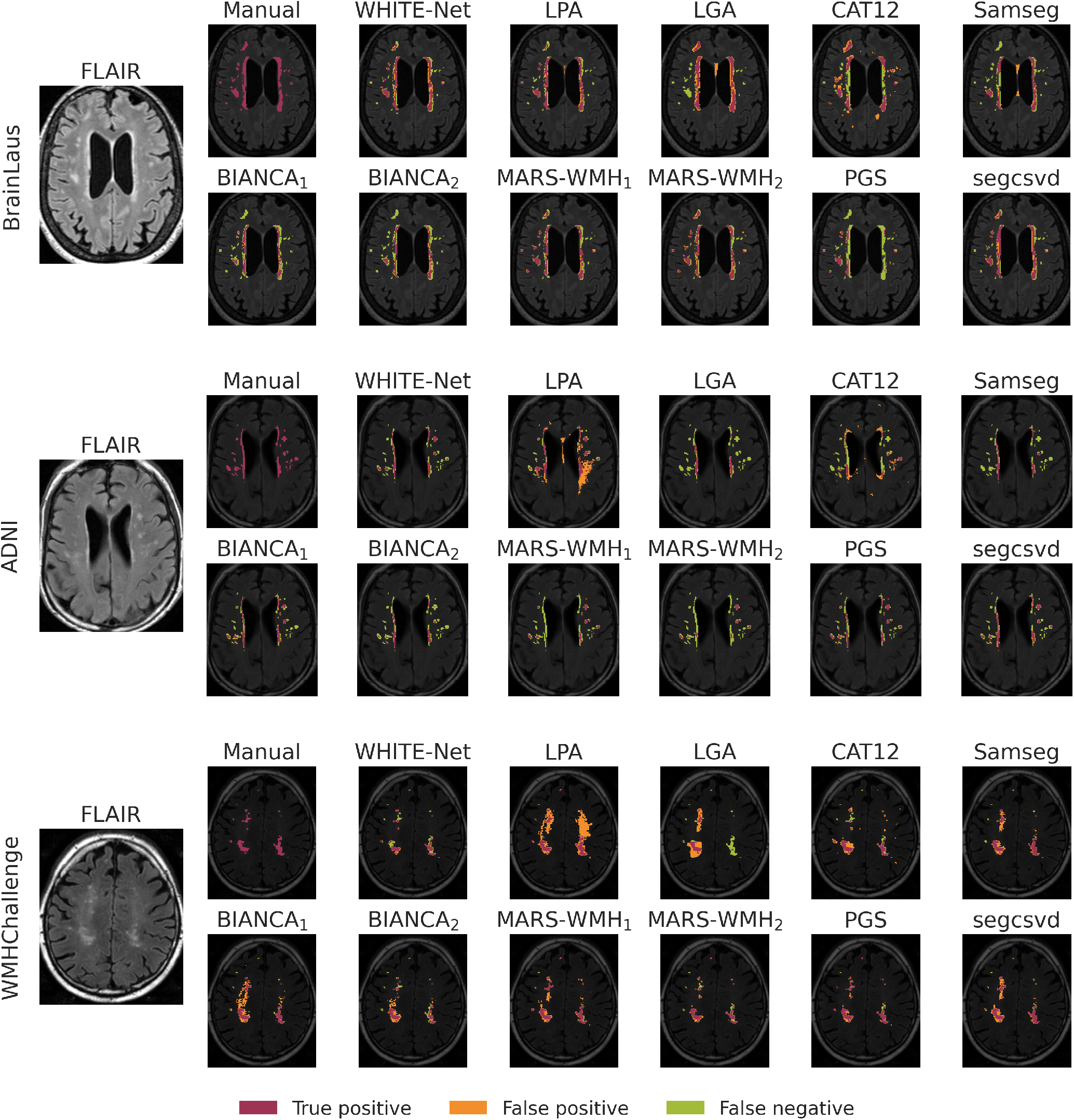
Examples of segmentation outputs across all methods, showing one representative brain from each dataset (BrainLaus, ADNI, and WMH Challenge). Pink, green, and orange indicate true-positive (TP), false-negative (FN) and, false-positive (FP) voxels, respectively. Supplementary Figure 5 shows additional BrainLaus examples across Fazekas scores 2, 4, and 6.

Overall, most methods capture the main lesion regions; however, noticeable differences are observed in lesion extent and false positive detections across datasets.

For the BrainLaus cohort, PGS underestimated the lesion burden, whilst in the ADNI LPA produced a high number of false positives compared with LGA, SamSeg, BIANCA, and MARS-WMH_2_ that failed to detect several lesions. In the WMH Challenge dataset, we observed a similar variability in segmentation quality with LPA and BIANCA largely overestimating lesion volume. WHITE-Net provided overall accurate segmentations.

### 3.4 WMH burden-stratified performance

Figure 4 illustrates the influence of lesion load on segmentation performance. Lesion load quartiles were defined based on log_10_(total lesion volume + 1), with lesion volume measured in mm^3^, pooled across datasets (Figure 4A). The distribution of subjects varied across datasets (Figure 4B), with BrainLaus enriched in low lesion loads, ADNI in moderate loads, and WMH Challenge in high lesion loads. Across methods, mean Dice increased with lesion burden (Figure 4C), a trend confirmed when stratifying by quartiles (Figure 4D), where performance was lowest in Q1 and highest in Q4. While consistent across methods, the magnitude of this effect varied. WHITE-Net and PGS showed stable performance across lesion loads, whereas MARS-WMH_2_ demonstrated greater sensitivity, with reduced Dice in low-lesion-load cases. Other methods were characterized by lower overall performance and higher variability. Variability across lesion load summarized by the standard deviation of Dice scores is shown in Figure 4E. WHITE-Net showed stable performance, PGS - dataset-dependent variability, and MARS-WMH_2_ higher variability in ADNI and WMH Challenge than in BrainLaus.

**Figure 4:**
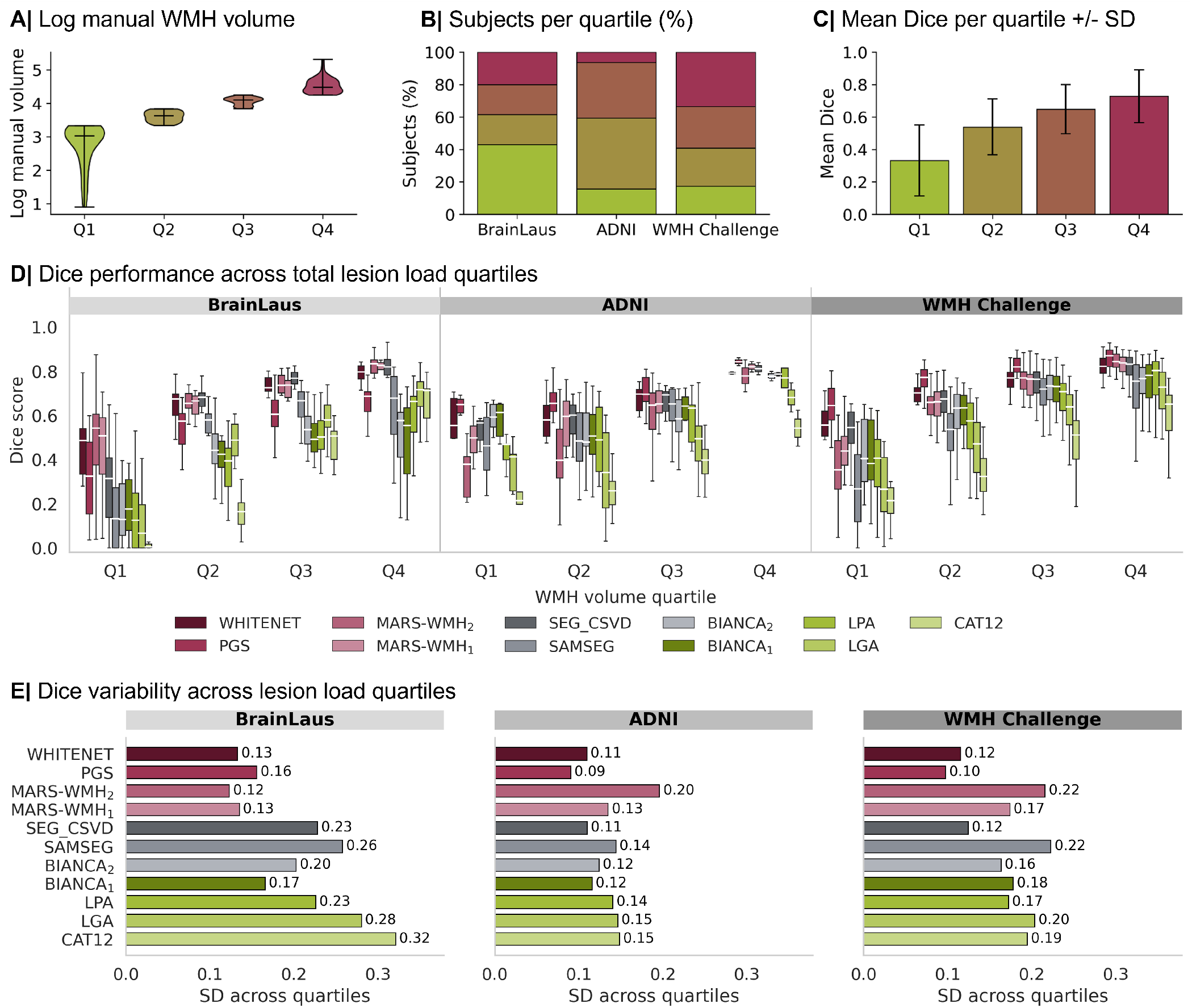
Influence of lesion load on segmentation performance. (A) Definition of lesion load quartiles based on log10-transformed total lesion volume [log_10_(volume + 1), mm^3^], pooled across the three datasets. (B) Percentage of subjects assigned to each WMH load quartile per dataset. (C) Overall mean Dice coefficient across all scans for each lesion load quartile. (D) Dice scores stratified by lesion load quartiles (Q1-Q4) for each method and dataset (BrainLaus, ADNI, WMH Challenge). Points denote median Dice scores, with horizontal bars indicating the interquartile range. Colors represent overall method performance, defined as the mean Dice across all subjects. (E) Standard deviation of median Dice scores across lesion load quartiles for each method, reflecting robustness to variability in lesion burden.

Segmentation performance was further evaluated in the BrainLaus dataset to assess method behavior across Fazekas score categories (Supplementary Figure 8A). WHITE-Net and MARS-WMH_2_ achieved the highest Dice scores across all Fazekas groups. PGS, SamSeg, BIANCA, LPA, and CAT12, showed markedly reduced performance in subjects with low Fazekas scores, highlighting the difficulty of segmenting low lesion-load cases. WHITE-Net demonstrated one of the lowest standard deviations in Dice scores, reflecting consistent performance across lesion burden categories, comparable to BIANCA_1_ (Supplementary Figure 8B). Unlike BIANCA_1_, WHITE-Net also achieved the highest overall Dice scores, demonstrating a favourable combination of accuracy and robustness across varying lesion burden.

### 3.5 Performance across lesion size

Figure 5 summarizes lesion-level detection performance across datasets calculated as described in Section 2.9.3. The total number of true positives (TP), false negatives (FN), and false positives (FP) per method is shown in Figure 5A, where FN and FP generally outnumber TP across methods. Lesions were stratified into quartiles based on log10-transformed total lesion volume [log10(volume + 1), mm^3^] (Figure 5B). Lesion size strongly influenced WMHs detection (Figure 5C): TP lesions were larger, FN lesions smaller, and FP lesions were predominantly small. This effect is further detailed by method and dataset (Figure 5D).

**Figure 5:**
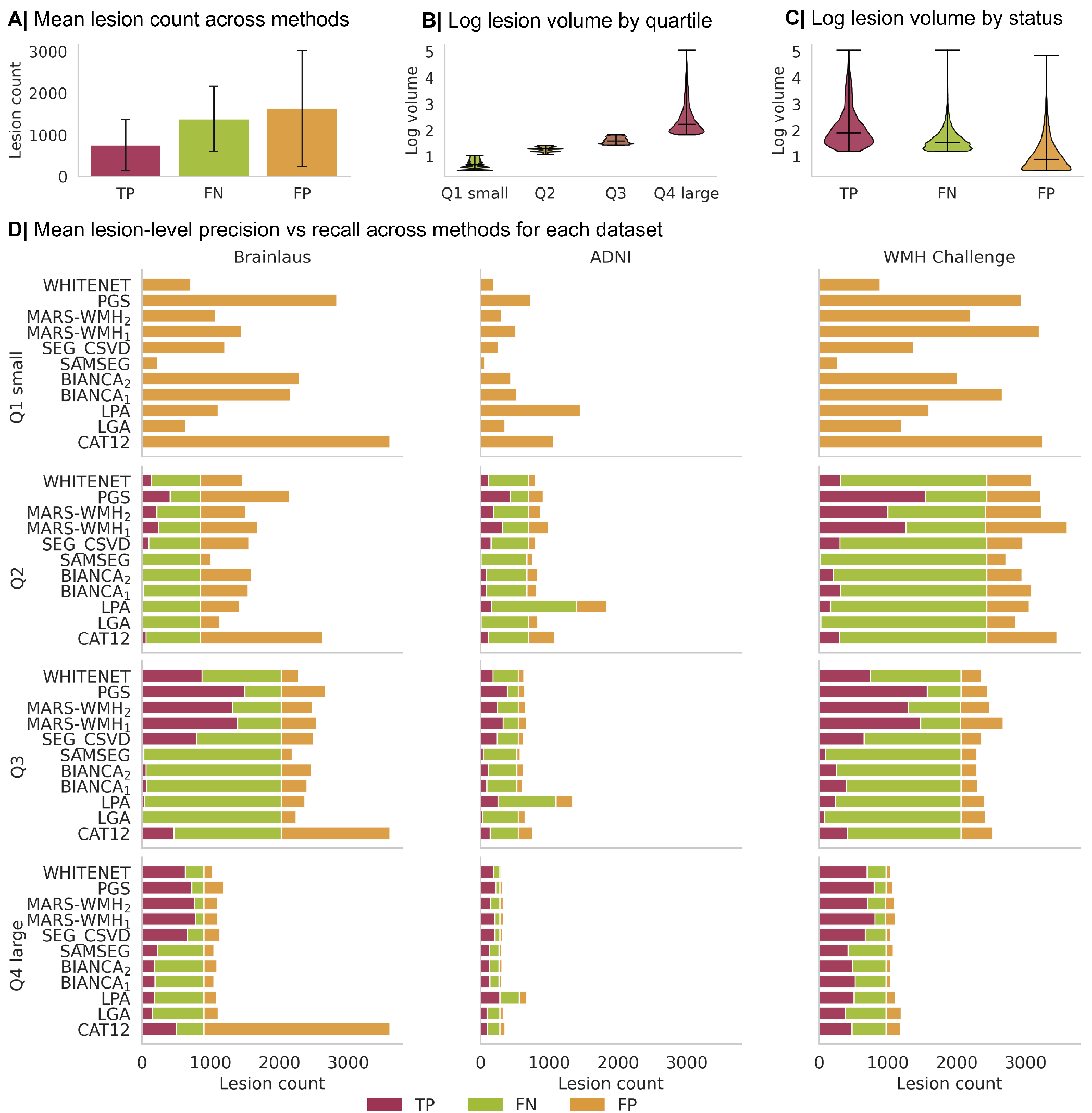
Lesion-level detection errors by lesion size and dataset. (A) Mean number of true-positive (TP), false-negative (FN), and false-positive (FP) lesions across methods. Error bars show standard deviation. (B) Distribution of lesion volumes across global lesion-size quartiles, shown on a log_10_(volume + 1) scale. (C) Distribution of lesion volumes by detection status. (D) Method-wise lesion counts stratified by lesion-size quartile and dataset. Horizontal stacked bars show TP, FN, and FP counts for each method in Brainlaus, ADNI, and WMH Challenge.

For the smallest lesions (Q1), nearly all WMH results were false positives. Several methods showed a substantial number of FPs, particularly in the WMH Challenge dataset, including PGS, MARS-WMH_2_, MARS-WMH_1_, BIANCA, and CAT12, while WHITE-Net and SAMSEG showed fewer FPs. For intermediate lesion sizes (Q2-Q3), BIANCA, LPA and LGA failed to detect a large proportion of TPs, whereas PGS and MARS-WMH_2_ achieved higher TP rates but for the cost of FP increases. For large lesions (Q4), FPs were rare across methods, with the exception of CAT12 in BrainLaus. WHITE-Net, PGS, MARS-WMH_2_, MARS-WMH_1_, segcsvd achieved a high number of TPs with a few FNs. In contrast, SAMSEG, BIANCA, LPA, LGA and CAT12 failed to detect a substantial number of large WMHs.

As a sensitivity analysis, we repeated the lesion-level comparison after applying the same removal of clusters <5 voxels to all methods (Supplementary Figure 6). This reduced FPs among the smallest lesions for some methods but did not substantially alter the overall pattern of results, with several methods continuing to show higher numbers of small FP lesions, particularly in the WMH Challenge dataset.

### 3.6 Performance stratified by clinical diagnosis

Method performance was evaluated across clinical subgroups in the ADNI dataset where participants were categorized as cognitively normal (NC) or with mild cognitive impairment (MCI). PGS and WHITE-Net achieved the highest Dice scores in both groups (Supplementary figure 8C). MARS-WMH_2_ showed greater inter-group performance variability, indicating reduced stability across clinical conditions. In contrast, while SamSeg, LGA, segcsvd, and CAT12 exhibited lower variability but consistently inferior overall Dice scores.

## 4 Discussion

### 4.1 Overall performance and generalizability

We present WHITE-Net, a deep learning tool for automated WMH segmentation trained on multisite MRI datasets encompassing substantial variability in scanner vendors, acquisition protocols, spatial resolution, and image characteristics. WHITE-Net achieves high segmentation accuracy and maintains robust performance across differing MRI data characteristics, lesion burden load and clinical diagnostic categories. WHITE-Net is open-source, requires no parameter tuning and offers fast execution times, making it readily deployable in large-scale epidemiological neuroimaging studies.

A key result is the consistent performance of WHITE-Net across datasets with heterogeneous imaging characteristics. While several methods achieved strong performance on specific datasets, WHITE-Net maintained competitive or top-level results across all cohorts ranking first on the monocentric BrainLaus dataset and second on both ADNI and WMH Challenge cohorts, reflecting robust generalizability across acquisition protocols. In contrast, methods such as PGS achieved high Dice scores on ADNI and WMH Challenge data but showed inferior performance on BrainLaus, suggesting greater sensitivity to dataset-specific characteristics. WHITE-Net demonstrated also good generalizability across varying lesion load, Fazekas scores, lesion size, and cognitive status.

The performance of existing methods should be interpreted in the context of their original training and validation settings. Competing approaches, including PGS (Park et al., 2021), segcsvd (Gibson et al., 2024), and the MARS-WMH_1_ and MARS-WMH_2_ models from MARS-WMH (Gesierich et al., 2025), were developed using different training datasets and acquisition settings. While these methods remained competitive, their performance varied across datasets and lesion burden, highlighting the influence of training and evaluation distributions on apparent performance. Therefore, our results compare available segmentation tools as implemented or pretrained, rather than their underlying architectures under matched training conditions.

### 4.2 Segmentation behavior and failure modes

Qualitative analysis further supports these findings, showing systematic differences in segmentation behavior across methods. While most methods successfully capture the main lesion regions, performance varied by dataset, reflecting sensitivity to site-specific imaging characteristics. This variability is also evident in the Dice scores, which fluctuate across sites for several methods and consistent with the well-established effects of training-to-testing distribution shifts (Matzkin et al., 2025). WHITE-Net demonstrated consistently high Dice scores and low cross-site variability in both ID and OOD settings with respect to the WHITE-Net training distribution, indicating robustness and generalizability. By contrast, other methods including PGS, segcsvd and MARS-WMH_2_, achieved competitive Dice scores overall but exhibited greater sensitivity to domain shifts: PGS and segcsvd showed increased variability within datasets classified as ID with respect to WHITE-Net with reduced accuracy in BrainLaus, while MARS-WMH_2_ exhibited more marked performance decrements under conditions classified as OOD with respect to WHITE-Net. These findings highlight that high average Dice scores are insufficient to characterize a method’s reliability, and that robustness to inter-site variability is a critical determinant of suitability for deployment across heterogeneous clinical and epidemiological MRI study settings.

Lesion-level analysis demonstrates that comparable Dice scores can arise from different error profiles. High recall methods such as PGS detect more lesions but at the cost of increased false positives, while more conservative approaches achieve higher precision at the expense of sensitivity. WHITE-Net favours precision over recall, yielding high detection reliability at the expense of reduced sensitivity to small or ambiguous lesions. This balance reflects an intentional design choice suited to the method’s primary use in research applications, where false positive detections risk including healthy tissue into downstream analyses. This is further supported by evidence that FLAIR-defined WMHs overestimate lesion spatial extent relative to histopathologically confirmed myelin loss (Haller et al., 2013). These findings highlight the importance of aligning segmentation behavior with the specific requirements of the intended application.

### 4.3 Influence of lesion burden

Segmentation performance was strongly influenced by total lesion burden, with performance declining in low lesion load across all investigated approaches. This effect was particularly pronounced for nn-Unet, whereas WHITE-Net showed more stable performance (Supplementary Figure 8). The ability to maintain reliable performance at low lesion burden is of particular relevance in studies of early stages of cerebrovascular disease or populations with low WMH burden, where accurate detection of small lesions is critical. The observed trend further highlights an inherent limitation of current segmentation approaches, which tend to perform better in cases with larger and confluent lesions.

Lesion size is a key determinant of segmentation performance, with smaller and sparser lesions more difficult to detect (He et al., 2025). Small lesions are frequently missed (FN), particularly by SAMSEG, BIANCA, LPA, LGA, and CAT12, whereas detection improves with increasing lesion size, with larger and confluent lesions more reliably identified as true positives. WHITE-Net takes a conservative approach to small lesion detection, yielding fewer false positives but missing a proportion of true small lesions. For larger lesions, however, its performance is comparable to other top-performing methods including PGS, MARS-WMH_1_, and MARS-WMH_2_.

Lesion location is an important additional determinant of automated segmentation performance, as WMHs are not uniformly distributed across white matter compartments (Wardlaw et al., 2015). They are more frequently observed in the DWM, while larger and confluent lesions predominate in the PVWM. Although certain regions such as periventricular watershed areas, are sites of lesion predilection, the precise spatial probability distribution of WMHs remains unclear (Caligiuri et al., 2015). Consistent with this, our results demonstrate reduced recall in the SWM, where lesions are generally smaller and harder to detect, while precision remained relatively homogeneous across compartments. The lower sensitivity in the SWM may be partly explained by the proximity to the cortical ribbon, where the low GM/WM contrast on FLAIR images renders tissue differentiation more challenging (Griffanti et al., 2016). These findings further emphasize the importance of anatomical stratification in the evaluation of WMH segmentation methods.

### 4.4 Robustness to scanner heterogeneity

Scanner variability had a notable impact on segmentation performance across methods (Dadar et al., 2017), reflecting heterogeneity in image contrast, noise levels, and spatial resolution (H. J. Kuijf et al., 2019). While WHITE-Net showed relatively stable performance across scanners, several competing methods exhibited substantial variability, suggesting reliance on scanner-specific intensity patterns or contrast characteristics that limit generalizability. The greater cross-scanner stability of WHITE-Net most likely reflects the diversity of its multisite training data, and reinforces the importance of training on heterogeneous datasets to achieve robust generalizability.

Complementing these observations, artifact-based perturbation analysis further highlighted the role of image acquisition characteristics in shaping model performance. MRI is particularly susceptible to motion artifacts (Zaitsev et al., 2015) and segmentation accuracy was strongly modulated by voxel anisotropy. Highly anisotropic scans showed increased sensitivity to motion and spatial resolution degradation, reflecting the compounded effects of reduced through-plane resolution and interpolation on image quality. WMH detection on isotropic image acquisitions were comparatively more robust, benefiting from spatially uniform information. Together, these findings reinforce that model robustness is not uniform across imaging conditions, but depends on both scanner-related variability and intrinsic acquisition properties, emphasizing the need for acquisition-aware evaluation and targeted robustness strategies. WHITE-Net’s performance on unprocessed images (no bias field correction) showed no significant impact on accuracy (Supplementary Figure 10), indicating that preprocessing is not required.

### 4.5 Design rationale and methodological contributions of WHITE-Net

WHITE-Net builds on the U-Net framework with design choices specifically targeting robustness to acquisition variability encountered in real-world neuroimaging data. While prior approaches have addressed related challenges through standardized acquisition and template-based alignment (Phuah et al., 2022), or anatomical priors (Liang et al., 2021), WHITE-Net instead targets domain shift directly through extensive artifact-driven data augmentation simulating variability in spatial resolution, motion, and intensity. This is complemented by a residual U-Net architecture stabilizing feature learning (Diakogiannis et al., 2020), a modified Dice loss enhancing sensitivity to small lesions, and an adaptive inference strategy adjusting decision thresholds to lesion burden (Sundaresan et al., 2019). These choices are supported by a highly heterogeneous training set comprising 22 acquisition parameter combinations across 13 sites and multiple scanner types, with evaluation spanning 11 acquisition configurations from 9 sites. By explicitly modeling variability at both training and inference stages in contrast to methods developed under more controlled conditions, WHITE-Net achieves consistent performance across datasets, particularly in out-of-distribution settings and at low lesion burden.

### 4.6 Computational efficiency and accessibility

WHITE-Net demonstrates high computational efficiency, with an execution time of only 28 seconds (Table 2). This fast processing speed is especially valuable for large cohort studies and clinical environments, where fast data processing is essential. Additionally, WHITE-Net requires no coding skills or parameter adjustments, making it highly accessible to users without technical expertise. Compared to other algorithms, WHITE-Net’s computational efficiency and simplicity make it a user-friendly tool.

### 4.7 Limitations

Several limitations should be acknowledged. First, although the datasets used in this study encompass a wide range of imaging conditions, they may not fully capture the diversity encountered in clinical practice, particularly across different pathologies, imaging protocols, or scanner configurations not represented in this work. As such, further validation on additional external datasets would be beneficial to confirm the generalizability of the findings. Second, the evaluation was conducted in a non-clinical computational anatomy setting. While this allows for controlled and systematic comparison of methods, it does not fully reflect real-world clinical workflows, where factors such as acquisition artifacts, comorbidities, or atypical lesion presentations may further impact performance (García-Lorenzo et al., 2013). Third, although Dice and lesion-level metrics provide complementary insights, they do not fully capture the clinical relevance of segmentation errors. For example, missing small lesions or incorrectly segmenting non-lesion tissue may have different implications depending on the downstream application. Future work could incorporate task-specific evaluation criteria, such as lesion-wise clinical relevance or longitudinal consistency.

An additional important consideration is the difference in training data exposure across methods. WHITE-Net was trained on multisite datasets with substantial variability in scanners and acquisition protocols, which likely contributes to its improved robustness compared to methods trained on more homogeneous data. This is consistent with prior work showing that training on heterogeneous, multisite data improves generalization and mitigates domain shift effects in medical imaging models (Zhang et al., 2020). While this may provide an advantage in the present comparison, it reflects an intentional design choice aligned with the goal of developing models that generalize across heterogeneous imaging conditions. In this context, the results highlight the value of training strategies that incorporate diverse data to improve robustness.

WMH annotations were derived from multiple datasets with different raters and protocols, and inter-rater reliability was not uniformly available. Although quality control procedures were applied, variability in manual segmentation remains, particularly for diffuse or low-burden lesions. This may introduce noise in both training and evaluation and should be considered when interpreting performance differences. At the same time, this heterogeneity reflects real-world clinical conditions and may contribute to improved robustness and generalizability. Future work should include larger datasets with standardized annotation protocols and comprehensive reliability assessments.

## Data Availability

Data of CoLaus|PsyCoLaus study used in this article cannot be fully shared as they contain potentially sensitive personal information on participants. According to the Ethics Committee for Research of the Canton of Vaud, sharing these data would be a violation of the Swiss legislation with respect to privacy protection. However, coded individual-level data that do not allow researchers to identify participants are available upon request to researchers who meet the criteria for data sharing of the CoLaus|PsyCoLaus Datacenter (CHUV, Lausanne, Switzerland). Any researcher affiliated to a public or private research institution who complies with the CoLaus|PsyCoLaus standards can submit a research
application to. Proposals will be evaluated by the Scientific Committee (SC) of the CoLaus|PsyCoLaus studies. Detailed instructions for gaining access to
the CoLaus|PsyCoLaus data used in this study are available at www.colaus-psycolaus.ch/professionals/how-to-collaborate/.
Data from WMH challenge are freely available at https://doi.org/10.34894/aecrsd (H. Kuijf et al., 2022).
Code for the segmentation tool described in this article, WHITE-Net, is available on GitHub (https://github.com/cathalacamille/WHITE-Net).

## 5 Data and Code Availability

Code for the segmentation tool described in this article, WHITE-Net, is available on GitHub (https://github.com/cathalacamille/WHITE-Net). The repository includes installation instructions, and guidelines for applying the model to new data. The current version of WHITE-Net requires only skull-stripped FLAIR MRI as input and can be applied directly without other mandatory preprocessing, although optional bias field correction may be used to help the model. Pretrained model weights are also provided to facilitate reproducibility and ease of use.

Data of CoLaus|PsyCoLaus study used in this article cannot be fully shared as they contain potentially sensitive personal information on participants. According to the Ethics Committee for Research of the Canton of Vaud, sharing these data would be a violation of the Swiss legislation with respect to privacy protection. However, coded individual-level data that do not allow researchers to identify participants are available upon request to researchers who meet the criteria for data sharing of the CoLaus|PsyCoLaus Datacenter (CHUV, Lausanne, Switzerland). Any researcher affiliated to a public or private research institution who complies with the CoLaus|PsyCoLaus standards can submit a research application to. Proposals will be evaluated by the Scientific Committee (SC) of the CoLaus|PsyCoLaus studies. Detailed instructions for gaining access to the CoLaus|PsyCoLaus data used in this study are available at www.colaus-psycolaus.ch/professionals/how-to-collaborate/. Data from WMH challenge are freely available at https://doi.org/10.34894/aecrsd (H. Kuijf et al., 2022). Data from ADNI and corresponding manual segmentations were provided by the team of Olivier Colliot (Vanderbecq et al., 2020).

## 6 Author Contribution

Data preprocessing was performed by AB, VR, and CC. Neural network design and model optimization were carried out by CC under the guidance of KF. FC performed quality control and clinically validated the annotations and Fazekas ratings. MH, QV and OC provided additional manually segmented datasets. AB and CC conducted the analyses and drafted the manuscript with contributions from all authors. AB, J-PT, and BD supervised the project.

## 8 Acknowledgments

BD is supported by the Swiss National Science Foundation (project grants Nr. 213595, 32003B_135679, 32003B_159780, 324730_192755 and CRSK-3_190185), ERA-NET NEURON JTC-2020 iSEE and ERA-NET NEURON JTC-2023 ELSA: BrainTree projects. LREN is very grateful to the Roger De Spoelberch and Partridge Foundations for their generous financial support. The research leading to these results has received funding from the French government under management of Agence Nationale de la Recherche as part of the “France 2030” program (reference ANR-23-IACL-0008, project PRAIRIE-PSAI) and as part of the “Investissements d’avenir” program (reference ANR-10-IAIHU-06, project Agence Nationale de la Recherche-10-IA Institut Hospitalo-Universitaire-6), from the European Union’s Horizon Europe Framework Programme (grant number 101136607, project CLARA), and from Inserm and the French Ministry of Health in the context of the MESSIDORE 2023 call operated by IReSP (reference AAP-2023-MSDR-341011).

## 9 Supplementary Material

**Supplementary Table 1:** Demographic characteristics of the BrainLaus sample stratified by Fazekas score.

| Fazekas | N | Age, mean ± SD | Age range | Female, n (%) |
| --- | --- | --- | --- | --- |
| 0 | 7 | 32.0 ± 10.7 | 22.2–54.6 | 7 (100.0%) |
| 1 | 21 | 46.5 ± 22.7 | 19.8–86.5 | 10 (50.0%) |
| 2 | 22 | 64.8 ± 12.8 | 31.1–81.3 | 9 (40.9%) |
| 3 | 22 | 72.6 ± 8.6 | 54.4–92.5 | 16 (72.7%) |
| 4 | 21 | 75.8 ± 6.0 | 65.4–85.5 | 9 (45.0%) |
| 5 | 20 | 75.0 ± 5.2 | 66.7–83.5 | 14 (70.0%) |
| 6 | 18 | 80.0 ± 7.0 | 66.7–92.8 | 13 (72.2%) |

**Supplementary Figure 1:**
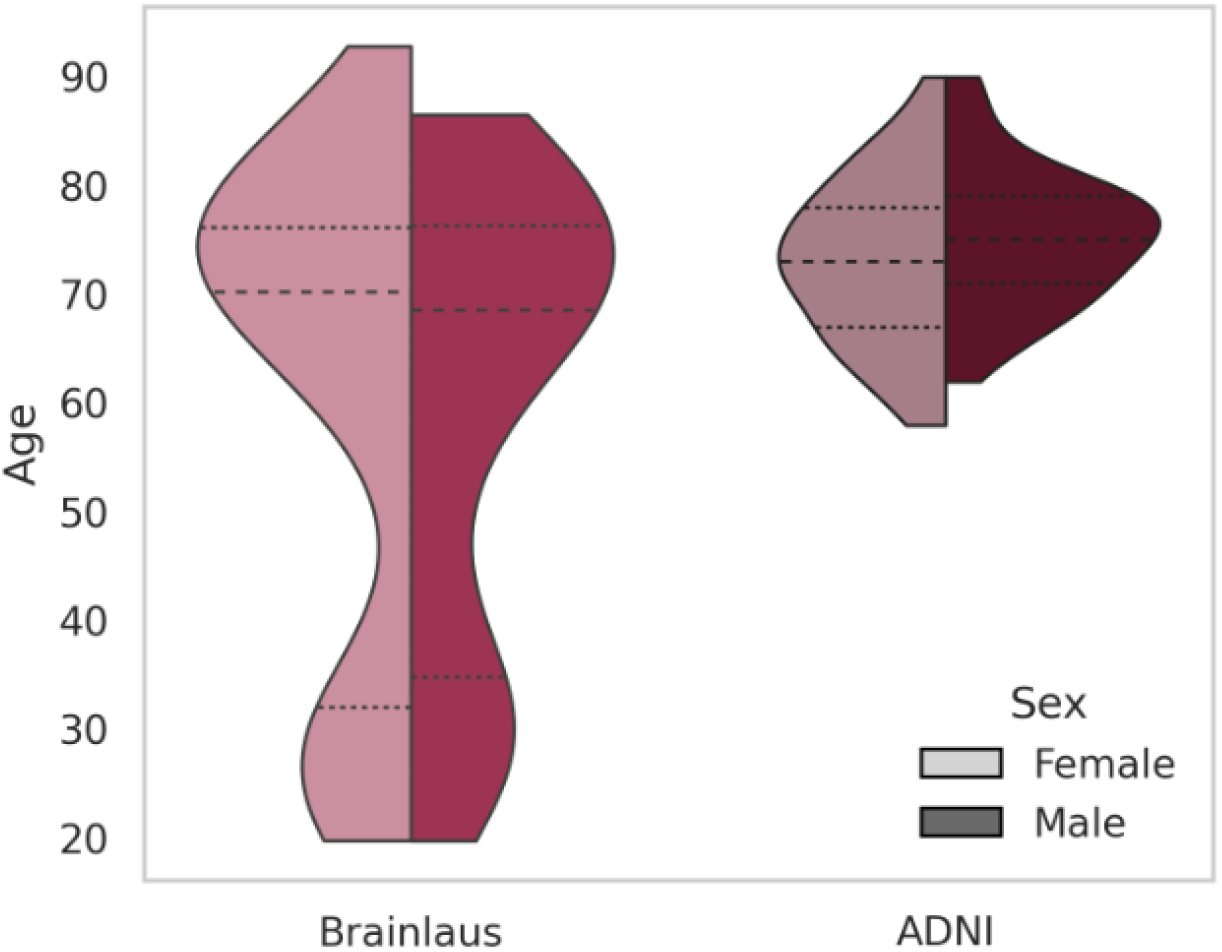
Age distributions stratified by sex for the BrainLaus and ADNI datasets.

We extracted a WM mask using Synthseg (Billot et al., 2023) and divided it into three compartments: deep white matter (DWM), periventricular white matter (PVWM) and superficial white matter (SWM). The SWM mask was defined as a 2mm thick outer contour of the total WM mask; the PVWM mask was obtained by dilating the cerebro-spinal fluid (CSF) mask by 8 mm; and the DWM mask was derived by subtracting the PVWM and SWM masks from the total WM mask (Supplementary Figure 9). Each lesion was assigned to the compartment with which it had maximal spatial overlap.

**Supplementary Figure 2:**
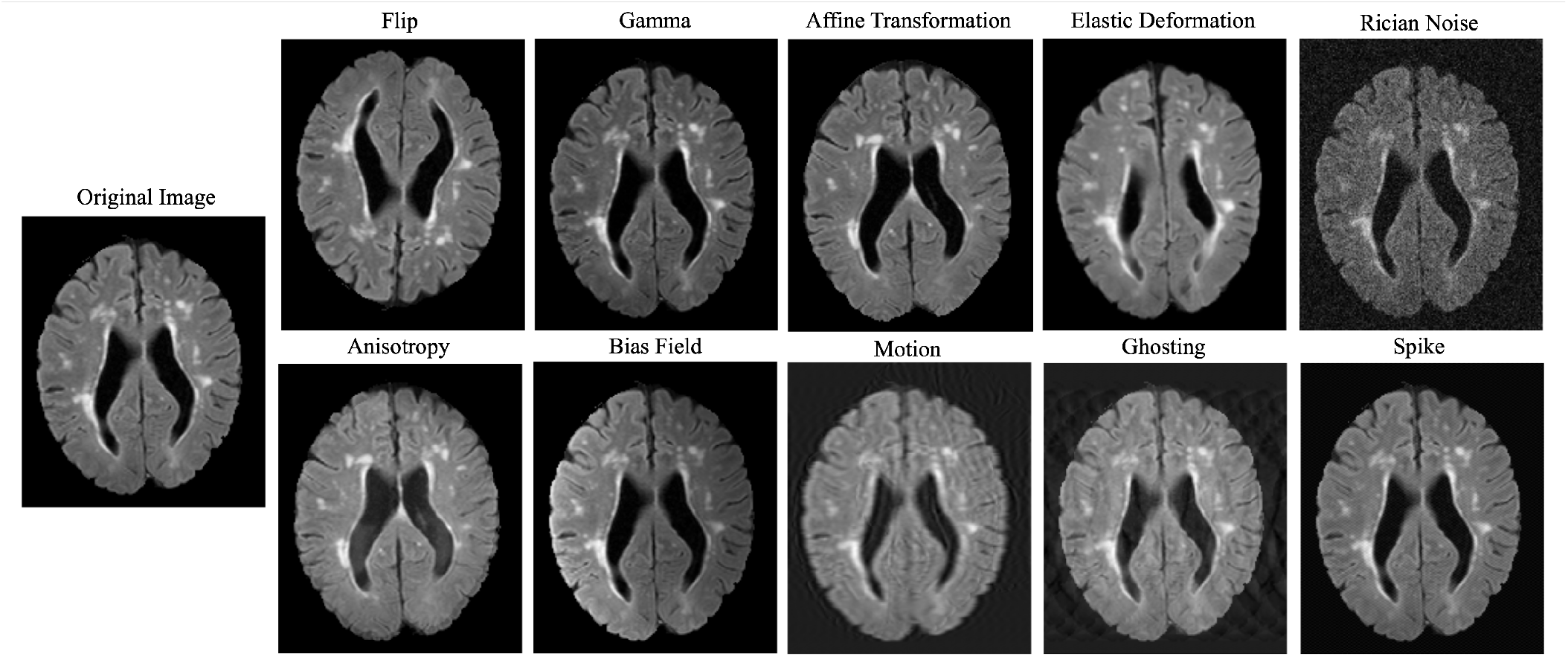
Original and augmented samples demonstrating the data augmentation strategy.

**Supplementary Figure 3:**
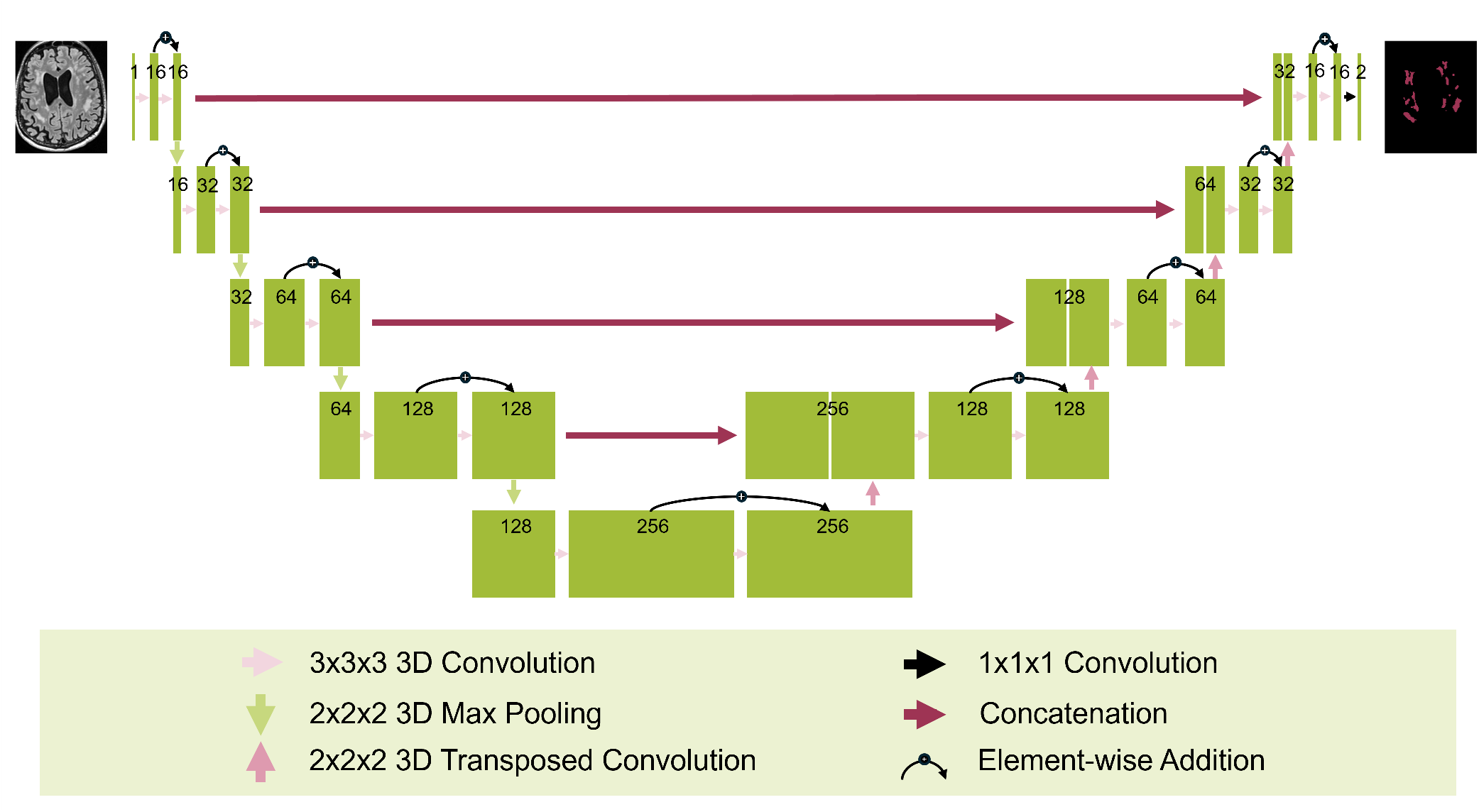
Overview of the ResUNet architecture. The encoder extracts features through convolution, ReLU, and max-pooling, with residual connections to improve gradient flow. The decoder restores spatial resolution via deconvolution and skip connections with encoder features.

**Supplementary Figure 4:**
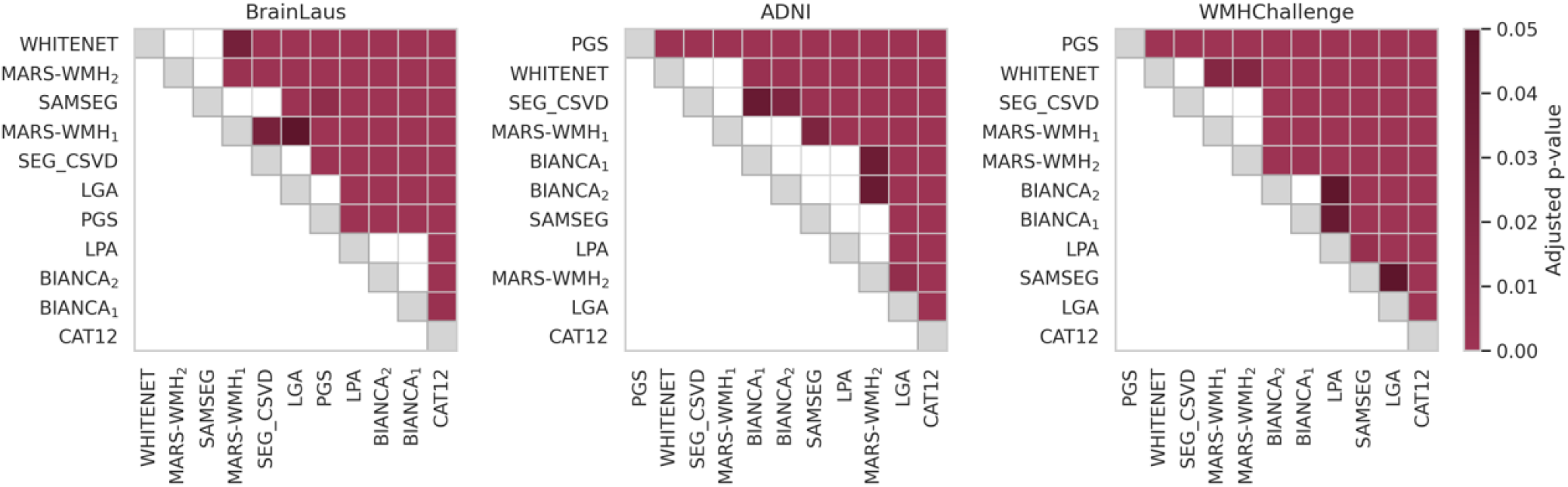
Pairwise Dice score comparisons between all methods, highlighting statistically significant differences. Colored cells indicate significant differences after FDR correction.

**Supplementary Figure 5:**
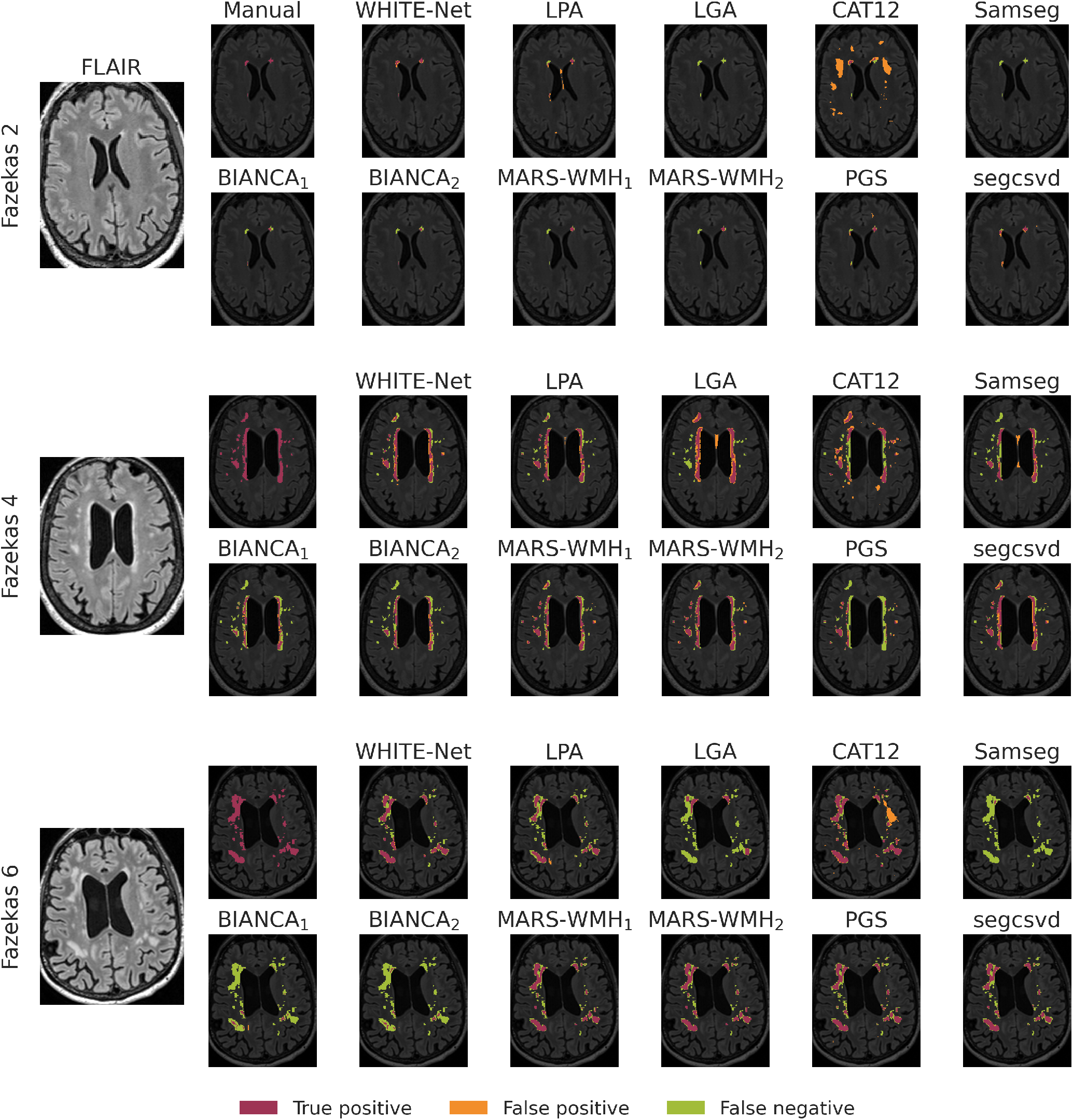
Examples of segmentation outputs across all methods in BrainLaus participants with different lesion loads, represented by Fazekas scores of 2, 4, and 6. Pink, green, and orange indicate true-positive (TP), false-negative (FN) and, false-positive (FP) voxels, respectively.

**Supplementary Figure 6:**
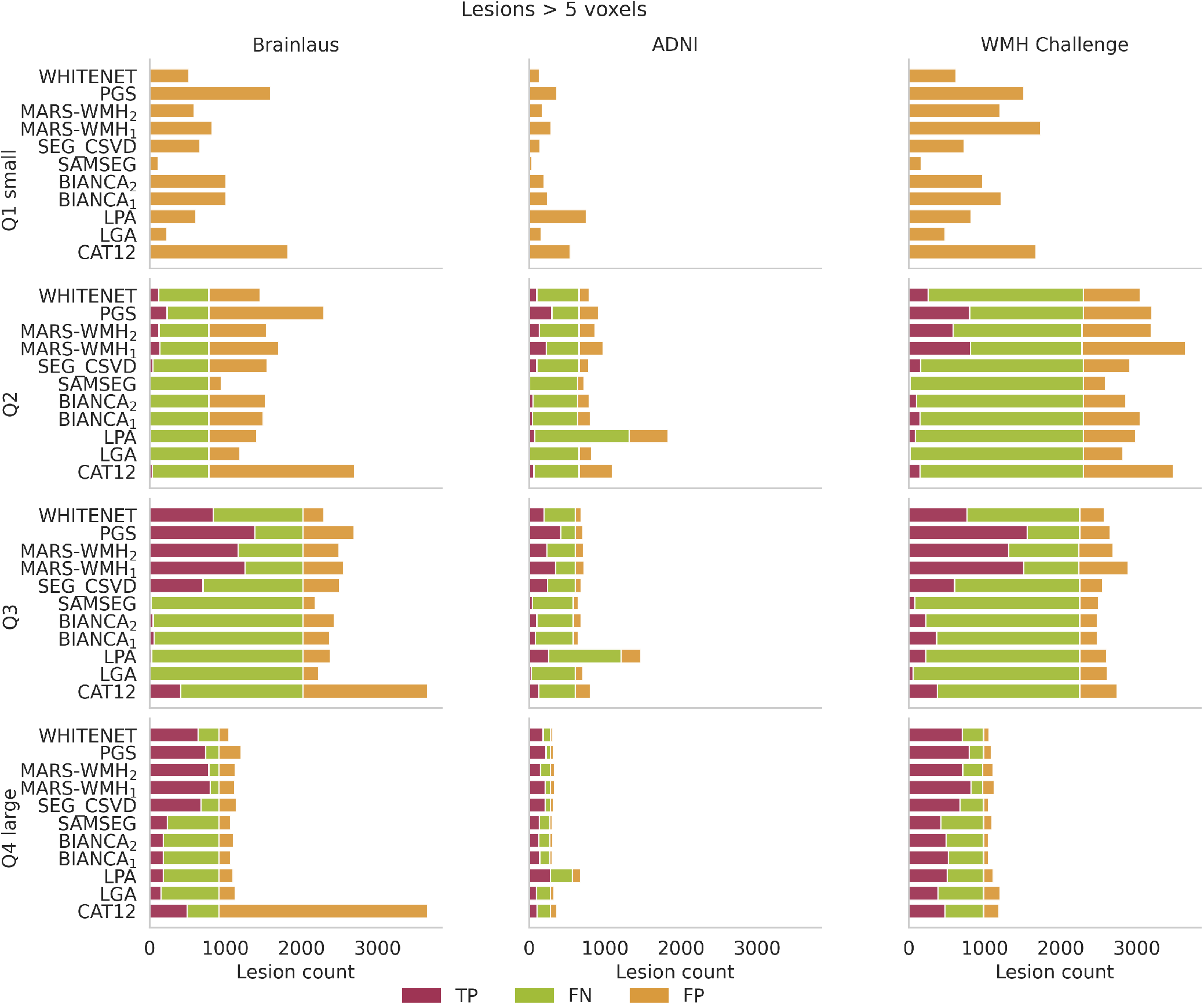
Lesion-level detection performance across lesion-size quartiles after applying a common post-processing step to all methods, removing predicted lesion clusters smaller than 5 voxels. Bars show the number of true-positive (TP), false-positive (FP), and false-negative (FN) lesions for each method and dataset.

**Supplementary Figure 7:**
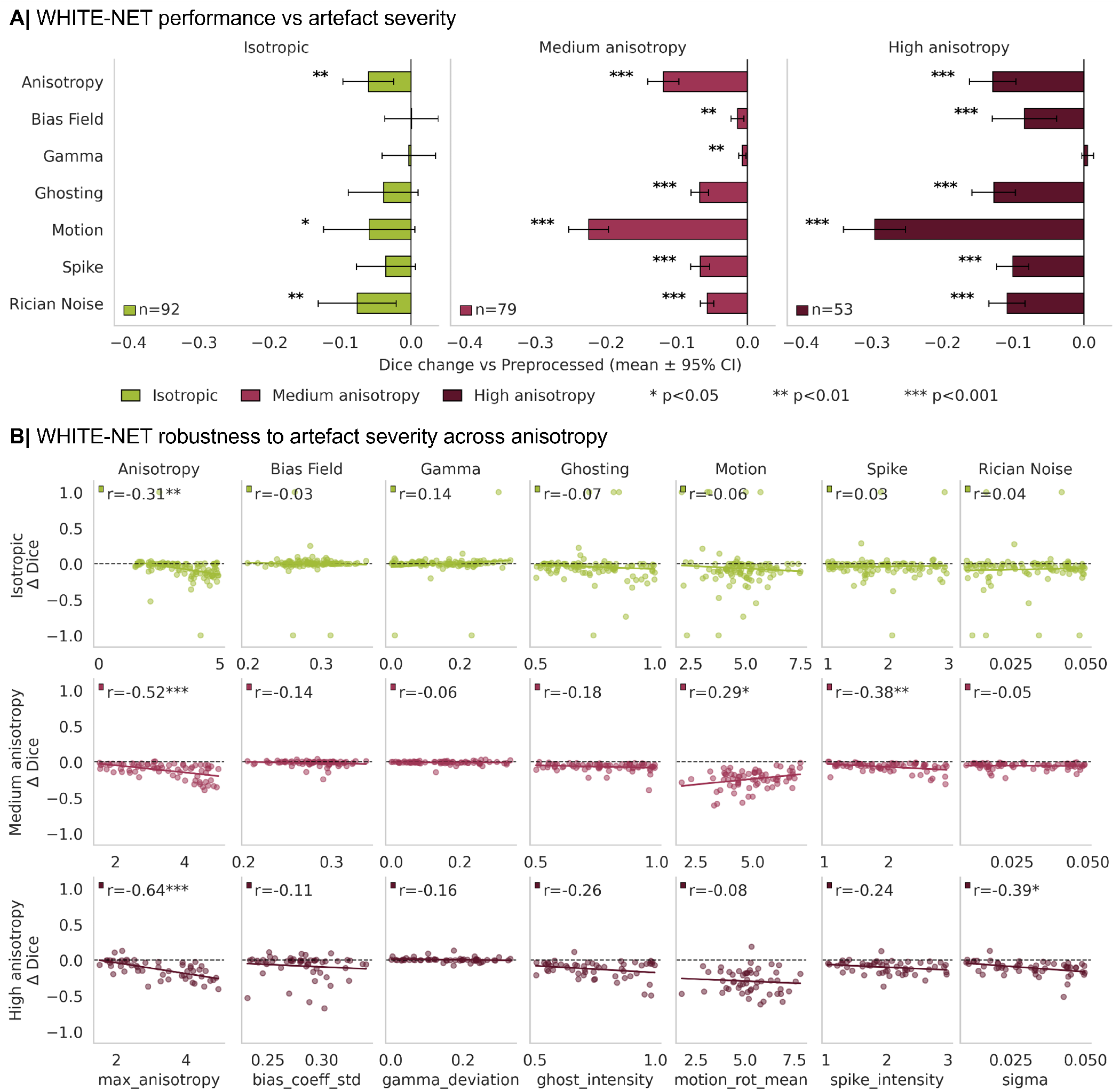
Impact of artificial artifact on Dice performance. (A) Change in WHITE-NET performance (ΔDice) for each simulated artifact (anisotropy, bias field, gamma, ghosting, motion, spike, and Rician noise), relative to the baseline Dice obtained from preprocessed images. Results are stratified into three subplots based on the original voxel anisotropy ratio: isotropic (green; voxel anisotropy ratio *≤* 1.01), moderately anisotropic (light pink; 1.01 *<* voxel anisotropy ratio *≤* 3), and highly anisotropic (dark pink; voxel anisotropy ratio *>* 3). Highly anisotropic scans exhibit greater sensitivity to artifact-induced degradation. (B) Relationship between artifact severity (randomly applied across test scans) and the corresponding reduction in Dice performance. Results are again stratified by the original scan anisotropy. Overall, higher artifact severity is associated with greater performance loss, particularly in anisotropic scans. Stars indicate statistically significant effects after FDR correction.

**Supplementary Figure 8:**
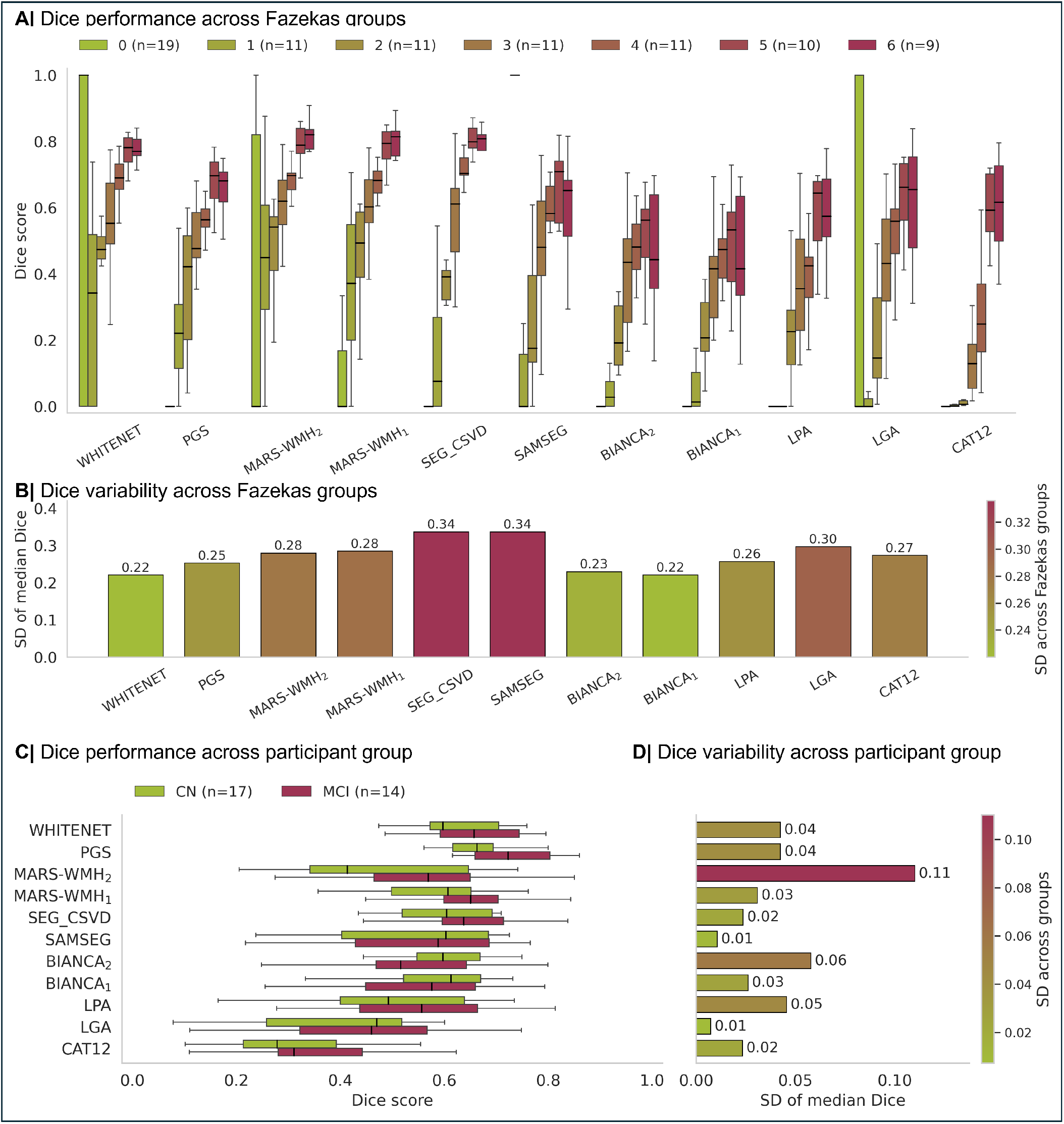
Influence of Fazekas score and participant group on segmentation performance. (A) Dice score stratified by Fazekas score (0–6) for each method in the BrainLaus dataset. (B) Standard deviation of median Dice scores across Fazekas scores for each method, quantifying robustness to lesion burden. (C) Median Dice score per method in ADNI dataset, comparing performance in controls and participants with mild cognitive impairment (MCI). (D) Standard deviation of median Dice scores across participant groups for each method, quantifying robustness to clinical status.

**Supplementary Figure 9:**
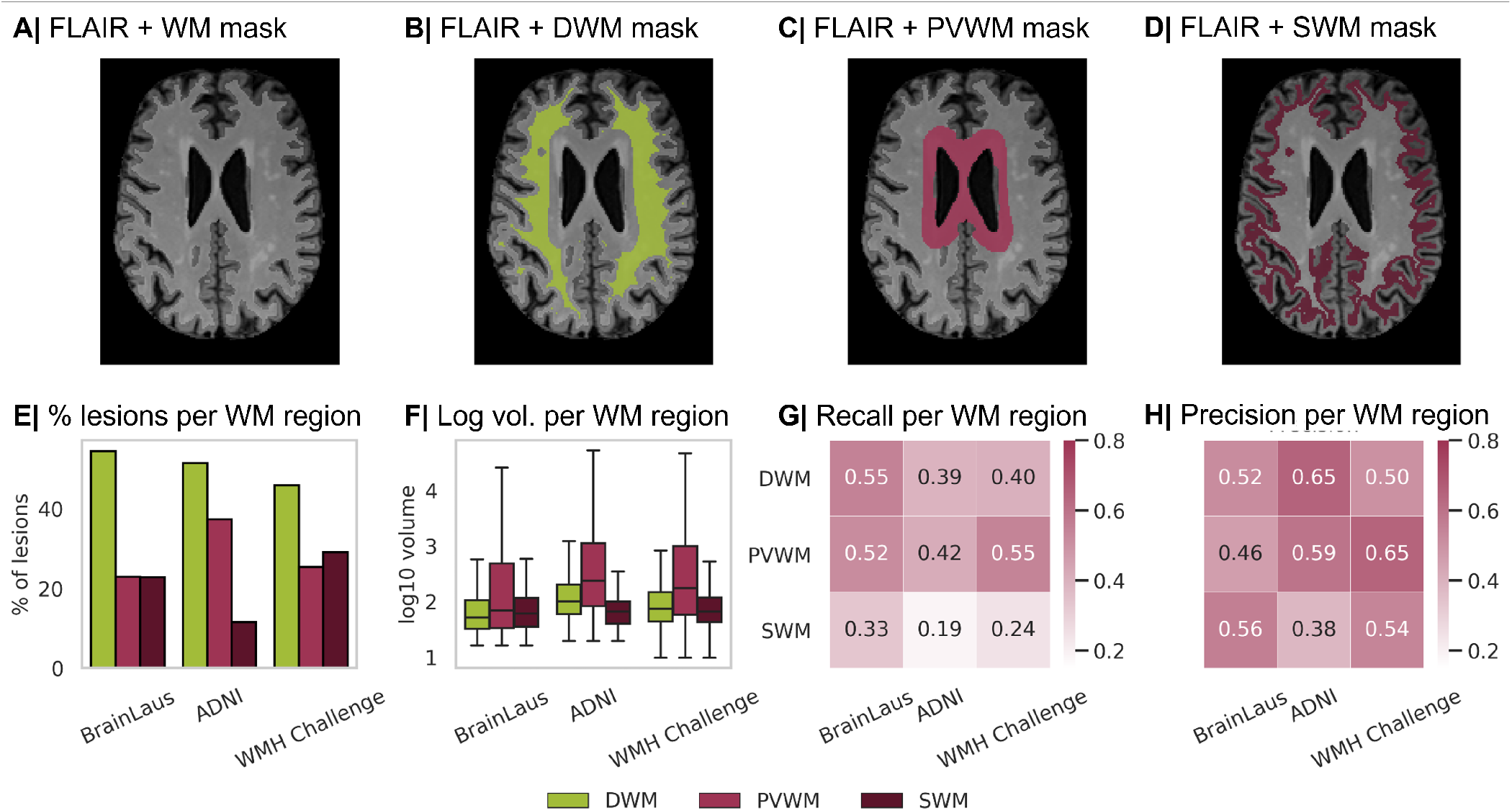
Model performance across WM regions. (A-D) Representative FLAIR MRI image with corresponding WM segmentation masks, including deep white matter (DWM), periventricular white matter (PVWM), and superficial white matter (SWM). (E) Distribution of lesions across WM regions, shown as the percentage of total lesions in each dataset. (F) Quantification of lesion burden expressed as log-transformed lesion volume for each WM compartment and dataset. Model performance metrics by WM region, including (G) recall and (H) precision.

**Supplementary Figure 10:**
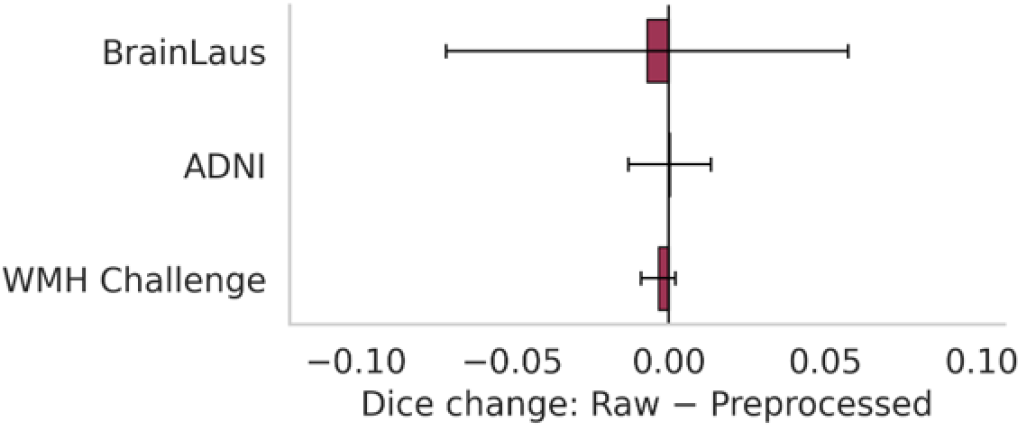
Mean difference in Dice score (raw - preprocessed) for WHITE-Net across datasets. Negative values indicate improved performance after preprocessing. Error bars represent 95% confidence intervals.

